# Multivariate genome-wide association analysis of quantitative reading skill and dyslexia improves gene discovery

**DOI:** 10.1101/2024.02.15.24302884

**Authors:** Hayley S. Mountford, Else Eising, Pierre Fontanillas, Adam Auton, 23andMe Research Team, Evan K. Irving-Pease, Catherine Doust, Timothy C. Bates, Nicholas G. Martin, Simon E. Fisher, Michelle Luciano

## Abstract

The ability to read is an important life skill and a major route to education. Individual differences in reading ability are influenced by genetic variation, with a heritability of 0.66 for word reading, estimated by twin studies. Until recently, genomic investigations were limited by modest sample size. Here we use a multivariate genome-wide association study (GWAS) method, MTAG, to leverage summary statistics from two independent GWAS efforts, boosting power for analyses of reading ability; GenLang meta-analysis of word reading (N = 27 180) and the 23andMe, Inc., study of dyslexia (N_cases_ = 51 800, N_controls_ = 1 087 070). We increase effective sample size to N = 102 082, representing the largest genetic study of reading ability, to date. We identified 35 independent genome-wide significant loci, including 7 regions not previously reported. Single-nucleotide polymorphism (SNP) based heritability was estimated at 24%. We observed clear positive genetic correlations with cognitive and educational measures. Gene-set analyses implicated neuronal synapses and proneural glioblastoma pathways, further supported by enrichment of neuronally expressed genes in the developing embryonic brain. Polygenic scores of our multivariate results predicted between 2.29-3.50% of variance in reading ability in an independent sample, the National Child Development Study cohort (N = 6 410). Polygenic adaptation was examined using a large panel of ancient genomes spanning the last ∼15k years. We did not find evidence of selection, suggesting that reading ability may not have been subject to recent selection pressure in Europeans. By combining existing datasets to improve statistical power, these results provide novel insights into the biology of reading.

## Introduction

Reading is a key academic skill and an important component of education. Difficulties with reading are associated with poorer life outcomes, lower socioeconomic status, and can greatly impact quality of life^1^. Unravelling the biological basis of this complex trait is essential in understanding the aetiology of reading, and why some people struggle with reading throughout their lives. A recent twin-based meta-analysis found that reading ability is highly heritable at 0.66^2, 3^, although prior estimates from classical twin models varied widely (0.03-0.83). Allelic variation in a number of genes have been associated with reading-related traits and dyslexia^4^, although with mixed support from replication studies. The largest genome-wide association study (GWAS) of quantitative reading skill^5^ (meta-analysis of 33 959 individuals from 19 cohorts) by the GenLang Consortium identified a single locus associated with word reading (rs11208009, P = 1.10×10^−8^) containing three candidate genes (*DOCK7*, *ANGPTL3* and *USP1*). Here, we improve statistical power using a much larger GWAS of dyslexia^4^ in a multivariate GWAS to improve gene discovery in investigations of reading ability.

Dyslexia, a common neurodevelopmental learning difference occurring in 5-10% of school age children^6^, is characterised by difficulties with accurate and/or fluent word reading, and poor spelling. Some diagnostic definitions of dyslexia extend to reduced performance on measures of verbal memory and processing speed, and/or emphasise a discrepancy between reading and other cognitive abilities^7^. Dyslexia tends to cluster within families^8^ and shows high heritability in twin-studies (0.4-0.6)^9^. Doust et al (2022)^4^ performed the largest GWAS of this trait to-date, using 23andMe, Inc. self-reported dyslexia diagnosis in 51 800 cases and 1 087 070 controls. Forty-two significantly associated regions were identified, including 27 not previously reported in studies of educational attainment or cognitive traits. Noteworthy for the current study, strong genetic correlations have been observed between dyslexia and quantitative measures of reading and spelling, ranging from −0.7 to −0.75 with CI 95% spanning −0.6 to −0.86^5^. These strong genetic correlations are consistent with the view that dyslexia is, to a large degree, representative of the low extreme of normal varying reading ability^10, 11^ rather than being a qualitatively distinct phenotype.

GWAS for dyslexia^4^ and quantitative reading skill^5^ clearly demonstrate the importance of large sample size to detect genetic effects that underlie reading traits. Eising et al’s (2022)^5^ GWAS meta-analysis of varied reading and spelling traits in up to 34 000 individuals made many important discoveries including revealing genetic correlations with other traits, significant SNP heritability (e.g., estimated at *h*^2^_snp_= 0.19 (SE = 0.02) for word reading), and clarifying the genetic structure among measures of reading, spelling, language, general cognitive ability, and educational attainment. However, it was relatively underpowered to detect single SNP variant associations that exceeded genome-wide thresholds. It is clear from studies of the genetics of reading^5, 12^, and also from other neurodevelopmental traits such as autism spectrum disorder (ASD)^13^ and attention deficit hyperactivity disorder (ADHD)^14^ with case sample sizes of 18 381 and 38 691, respectively, that analysing large sample sizes is key to improving resolution of associated variants. For neurodevelopmental measures of literacy collected in childhood, it has historically been challenging to gather sufficient sample size, primarily due to the fact that reading skill is not perceived as medically relevant. Meta-analysis approaches are appropriate for addressing this issue.

One of the alternative methods to collecting and phenotyping new cohorts for a trait of interest, multi-trait analysis of GWAS approach (MTAG)^15^ which uses the shared genetic architecture of related phenotypes (multivariate) to increase gene discovery power. For example, Grove and colleagues^13^ used MTAG to increase their ASD GWAS power by adding GWAS for schizophrenia, educational attainment, and major depression. This showed stronger evidence for previously reported regions, and seven novel regions shared with educational attainment or depression. More recently, multivariate analyses were used across five psychiatric traits (ASD, ADHD, bipolar disorder, schizophrenia, and depression)^16^. Again, this increased the number of associated loci identified for each individual trait, particularly bipolar disorder, which increased from 8 genome-wide significant loci to 54. In addition to statistical power gain, this multivariate associated approach permits to delineate the shared and separate genetic profiles of related conditions.

Given the strong genetic correlation between dyslexia and word-reading skills^4^, we applied the multivariate method to boost sample size of the word reading GWAS and identify novel associated loci. We note that minimal sample overlap is expected, given that the GWAS of dyslexia includes a single sample of adult predominantly US-based 23andMe, Inc. research participants and the GWAS of word reading is a meta-analysis of cohorts of children and young adults with a minority from the US. Moreover, MTAG uses bivariate LD score regression to control for sample overlap between cohorts.

By increasing effective sample size through multivariate GWAS, we aimed to achieve the following. Firstly, improved power to detect novel loci associated with reading, uncovering biological pathways at the gene and functional level, and linking to biological mechanisms that underlie reading ability through the availability of developmental brain gene-expression datasets^17^. Secondly, obtaining sufficient power to generate reliable polygenic scores to predict reading variation in independent cohorts. Doust et al^4^ generated a polygenic score from the dyslexia GWAS which significantly predicted variance in reading performance measures in independent samples, for example, accounting for 3.6% in nonword reading in the Brisbane Adolescent Cohort and 5.6% of variance in word recognition in a US cohort enriched for individuals with reading difficulties (Colorado Learning Disabilities Research Centre)^4^. Finally, we used a novel method to determine if the polygenic scores for word reading showed evidence of selection in the past 15 000 years^18, 19^.

## Materials / Subjects and Methods

### Ethics approval statement

The study made use of existing data sets and was granted ethical approval by the University of Edinburgh School of Philosophy, Psychology and Language Sciences research ethics committee (PPLSREC 29-1819/8).

### Multivariate GWAS

GWAS summary statistics for quantitative measures of word reading were available in the GenLang meta-analysis of 27 180 (male = 13 874, female = 13 202, no information = 104) participants of European ancestry only across 18 studies^5^. Participants were children or young adults ranging from 5-26 years^5^. Measures depended on the contributing cohort and are detailed in Eising *et al*^5^, with the Test of Word Reading Efficiency (TOWRE) and Wide Range Achievement Test (WRAT) most common. Summary statistics without genomic-control correction were used.

Summary statistics for self-reported dyslexia diagnosis (23andMe, Inc.) were reported in Doust *et al* (2022)^4^. Participants responded “Yes” to “have you been diagnosed with dyslexia” (N_cases_ = 51 800, female = 30 287, male = 21 513), and N_controls_ = 1 087 070 responded “No” (female = 641 016, male = 446 054). All participants were over 18 years of age (mean 49.6 (cases) and 51.7 years (controls)). Participants with non-European ancestry were excluded prior to analysis. Summary statistics without genomic-control correction were used^4^.

Both sets of summary statistics were annotated with rsIDs (build hg19), formatted for MTAG, and Z scores were calculated in R. Variants with imputation quality <0.8 or minor allele frequency of <0.01 were excluded prior to analysis. Multivariate GWAS was performed with MTAG^15^ using default settings and false discovery rate (FDR) calculation. Associations were visualised using ggplot2^20^.

Individual regions were visualised using LocusZoom (http://locuszoom.org). FUMA version v1.5.0 was used to annotate the associated regions that met the genome-wide significance threshold^21^ (P ≤5×10^−8^, *R*^2^ <0.6, and <250kb maximum distance between LD blocks to merge into one genomic locus).

### Heritability and Genetic Correlation

SNP-based heritability (*h*^2^_snp_) was estimated using LDSC v1.0.1^22^. European LD reference panel was obtained from https://alkesgroup.broadinstitute.org/LDSCORE. GWAS summary statistics were obtained from the Complex-Traits Virtual Genetics Lab (CTG-VL) platform (https://vl.genoma.io), except for ASD^13^ and ADHD^14^ which were downloaded from the Psychiatric Genomics Consortium repository (https://www.med.unc.edu/pgc/download-results). Genetic correlations were performed using LD-Score v1.0.1 within the CTG-VL platform and considered significant at a Bonferroni corrected threshold of P ≤ 3.478×10^−5^ from 1438 tests.

Due to the difference in sample size between the Dyslexia and GenLang univariate summary statistics, we cross-checked genetic correlations between each set of summary statistics and the MTAG output to check that the contributions were approximately equivalent, assessed by false discovery rate (FDR).

### Gene-based and Gene-set Analysis

Gene-based associations were calculated using MAGMA v1.08^23^ using SNP2GENE within the FUMA interface (https://fuma.ctglab.nl/) for 18 842 genes and were considered significant at a Bonferroni corrected threshold of P ≤ 2.65×10^−6^. Gene-set analyses of biological pathways defined by gene ontology (GO) pathways and curated gene-sets were examined using MAGMA. GO terms and gene-sets containing fewer than 20 genes were excluded from analysis, so a total of 9 113 biological pathways were tested and a Bonferroni corrected threshold of P ≤ 5.49×10^−6^ was applied.

### Functional Mapping, Annotation and Partitioned Heritability

Fine mapping and annotations were performed using the Variant Effect Predictor (VEP) online tool (http://grch37.ensembl.org/) on the list of candidate SNPs present with R^2^ ≤0.6 with an independent significant SNP generated by FUMA, and includes tagged SNPs extracted from the 1000 genomes reference panel. Variants were considered potentially damaging if they were annotated as probably damaging by PolyPhen2 and deleterious by SIFT. Expression QTL analysis was performed using PsychENCODE eQTLs, BRAINEAC, eQTLcatalogue BrainSeq, and GTEx v8 Brain databases within FUMA.

MAGMA, within FUMA, was used to test for enrichment of tissue-specific annotations. For this, we used bulk RNA-seq expression profiles from 53 tissue types from GTEx v8, and BrainSpan RNA-sequencing from 29 ages spanning 11 developmental stages.

To interrogate cell- and region-specific resolution, we accessed single-cell RNA-seq (scRNA) data via the Cell Type function within FUMA. Expression data from human embryonic ventral mid-brain (6-11 weeks post gestation) (GSE76381), human embryonic prefrontal cortex (8-26 weeks post gestation) (GSE104276), and human adult and fetal cortex (GSE67835) datasets were tested.

We partitioned SNP heritability using stratified LDSC, as described by Finucane *et al*^24^, to determine if significantly more SNPs clustered within tissue-specific chromatin modification patterns than expected by chance, based on the proportion of SNPs that map within these types of genomic regions. Annotations were based on data from the Roadmap Epigenomics project and Enhancing GTEx project (ENTEx). LD scores, regression weights and European allele frequencies were obtained from https://alkesgroup.broadinstitute.org/LDSCORE.

### Polygenic Score Analysis

Word-reading polygenic scores were calculated for SNPs based on increasing P value thresholds spanning the full range. Scores were calculated for the National Child Development Study (NCDS); a large UK birth cohort study born in 1958^25^, with extensive genetic and longitudinal reading measures as described in Bridges *et al*^26^. Individuals genotyped on one of the two lowest resolution arrays (Illumina 15k Custom chip and Affymetrix 500k) were excluded prior to analysis due to lack of available data from chromosome X, resulting in a cohort of N = 6 410 with imputed genotypes. Six measures of functional reading were used, consisting of composite measures at ages 7, 11, 16 and across all time points, and binary measures of difficulties at ages 23 and 33^26^. Polygenic scores were estimated using PRSice2 v2.3.5^27^, and sex, array and 10 principal components were used as covariates, as previously described^26^.

### Polygenic Selection Analysis

We sought to identify evidence of polygenic selection for reading ability using a large panel of 1015 imputed ancient genomes, sampled from across West Eurasia^19, 28^. This dataset represents a dense transect of ancient individuals sampled over the last 15k years with local ancestry contributing to present day Europeans. We ascertained statistically independent SNPs associated with reading ability by filtering our genome-wide summary statistics to only retain positions imputed with high confidence in the ancient dataset. LD-clumping was performed using Plink 1.90b4 with a window size of 250kb, maximum *R*^2^ threshold of 0.05, and maximum P ≤ 5×10^−8^ using the 1000 Genomes Project phase 3 European populations (GBR, FIN, TSI) as the reference panel. We then inferred allele frequency trajectories and selection coefficients using CLUES^29^ and exported the posterior likelihood densities. Finally, we modelled the polygenic selection gradient for reading ability with PALM^30^ using imputed ancestral data generated and method described by Barrie *et al*^18^.

## Results

### Multivariate GWAS of Reading Ability

The genetic correlation between the univariate summary statistics of word reading and dyslexia (Europeans only, without GC correction) was −0.71 (SE = 0.05, Z = −15.06), and indicative of a high degree of shared genetic aetiology enabling multivariate GWAS analysis with MTAG^15^. The analysis produced an equivalent sample size of 102 082 individuals for reading ability and used 5 449 985 DNA variants that were shared between the two sets of univariate summary statistics. This provided 87% power to detect additive trait variance of up to 0.04% (N = 102 082, α = 5×10^−8^). We identified 35 genome-wide significant (P ≤ 5×10^−8^) independent loci (*R*^2^ <0.6, and <250kb maximum distance between LD blocks to merge into one genomic locus) (figure 1a), containing 86 independent significant SNPs, independent from each other at an *R*^2^ of 0.1. Twenty-six of these regions were previously reported in the dyslexia GWAS^4^, 6 regions can be considered novel but were present in the uncorrected dyslexia summary statistics, and 1 region is novel (figure 1, table 1). The region previously reported as significant in the GenLang word reading meta-analysis^5^ was not associated in the present study. Quantile-Quantile (Q-Q) plots indicated (supplementary figure 1) that the data were appropriately controlled for population stratification, as markers showing low association with word reading did not deviate from the expected quantile. Similarly, LDSC lambda genomic control statistics (1.28), intercept (0.84, SE = 0.01) and ratio (<0) support an absence of genomic inflation. The maximum FDR for MTAG was moderate at 0.06, indicating that 6% of associated regions may be false positives. Constituent univariate GWAS χ^2^ statistics were calculated by MTAG for dyslexia (χ^2^ = 1.66) and GenLang word reading (χ^2^ = 1.08), and multivariate GWAS (χ^2^ = 1.35). Individual LocusZoom plots for each region are presented in supplementary figure 2:1-35. Summary statistics for SNPs reaching suggestive significance (P ≤ 1×10^−5^) are presented in supplementary table 1.

**Figure 1:**
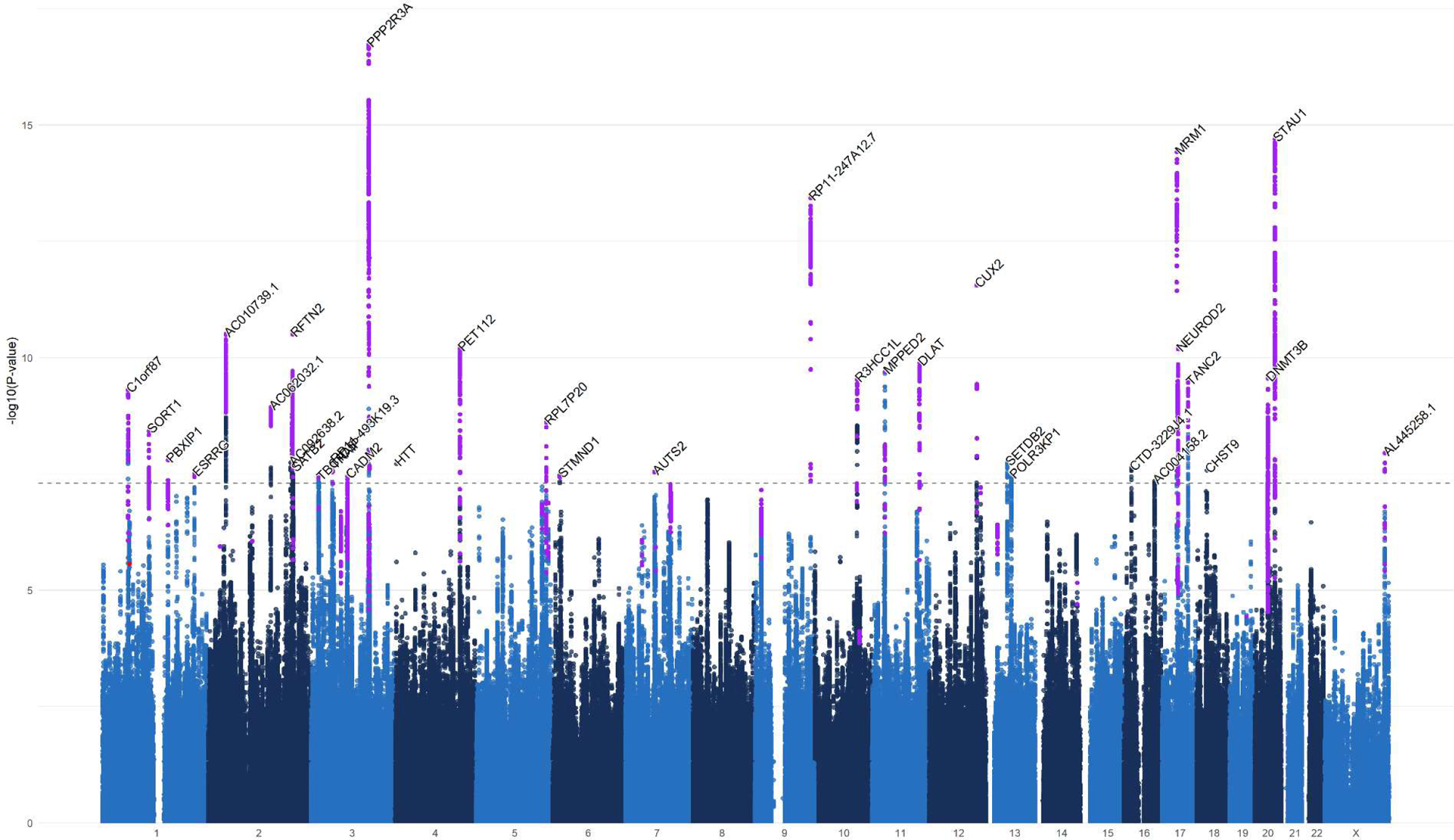
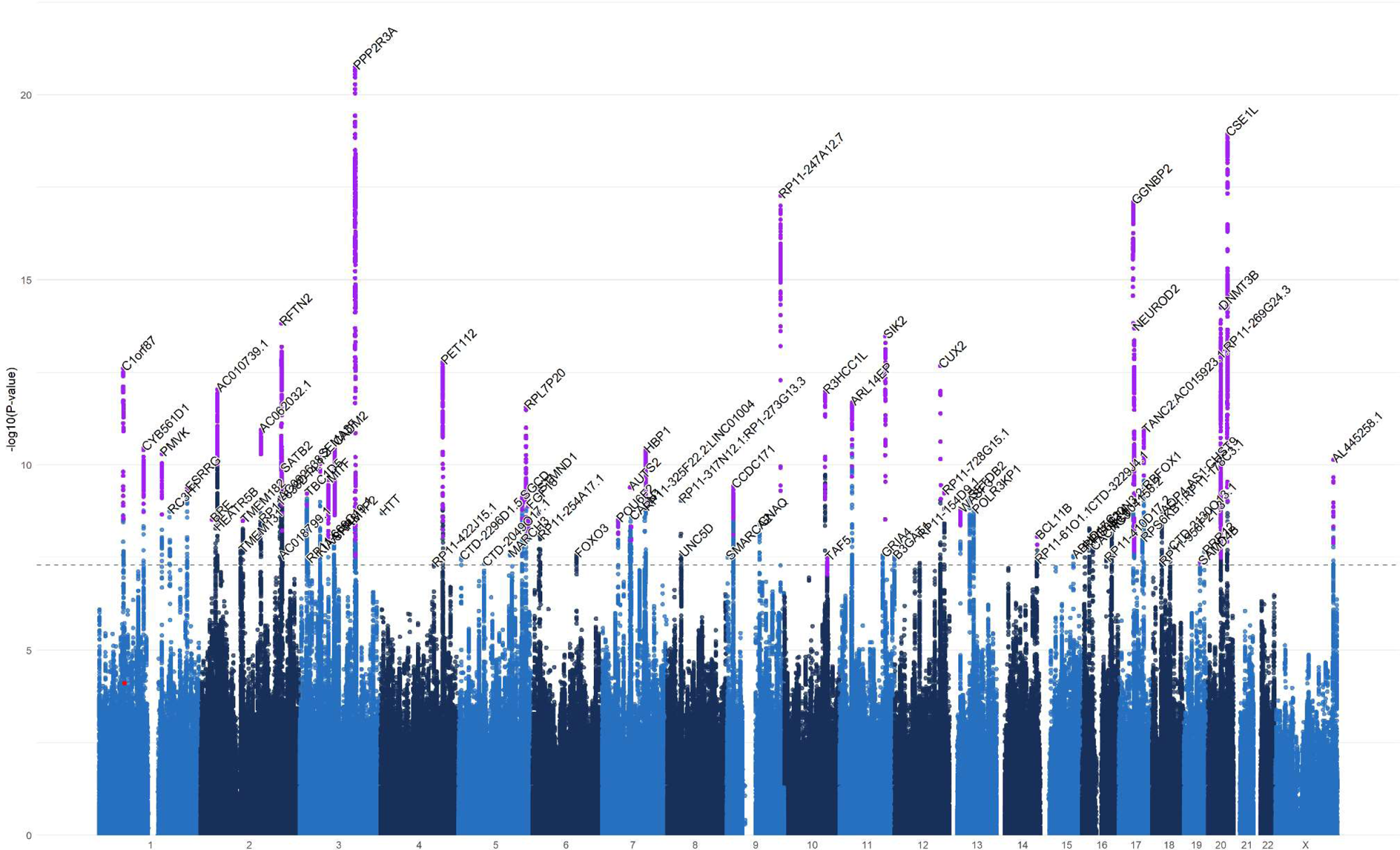
Manhattan plot of the multivariate GWAS of. a) reading ability and b) dyslexia. The y axis indicates the −log_10_ P value for association. The threshold for genome-wide significance (P< 5×10^−8^) is represented by a dashed grey line. Significant loci that were previously reported in the GenLang word reading GWAS are represented in red, and those reported in the dyslexia GWAS are shown in purple.

**Table 1:**
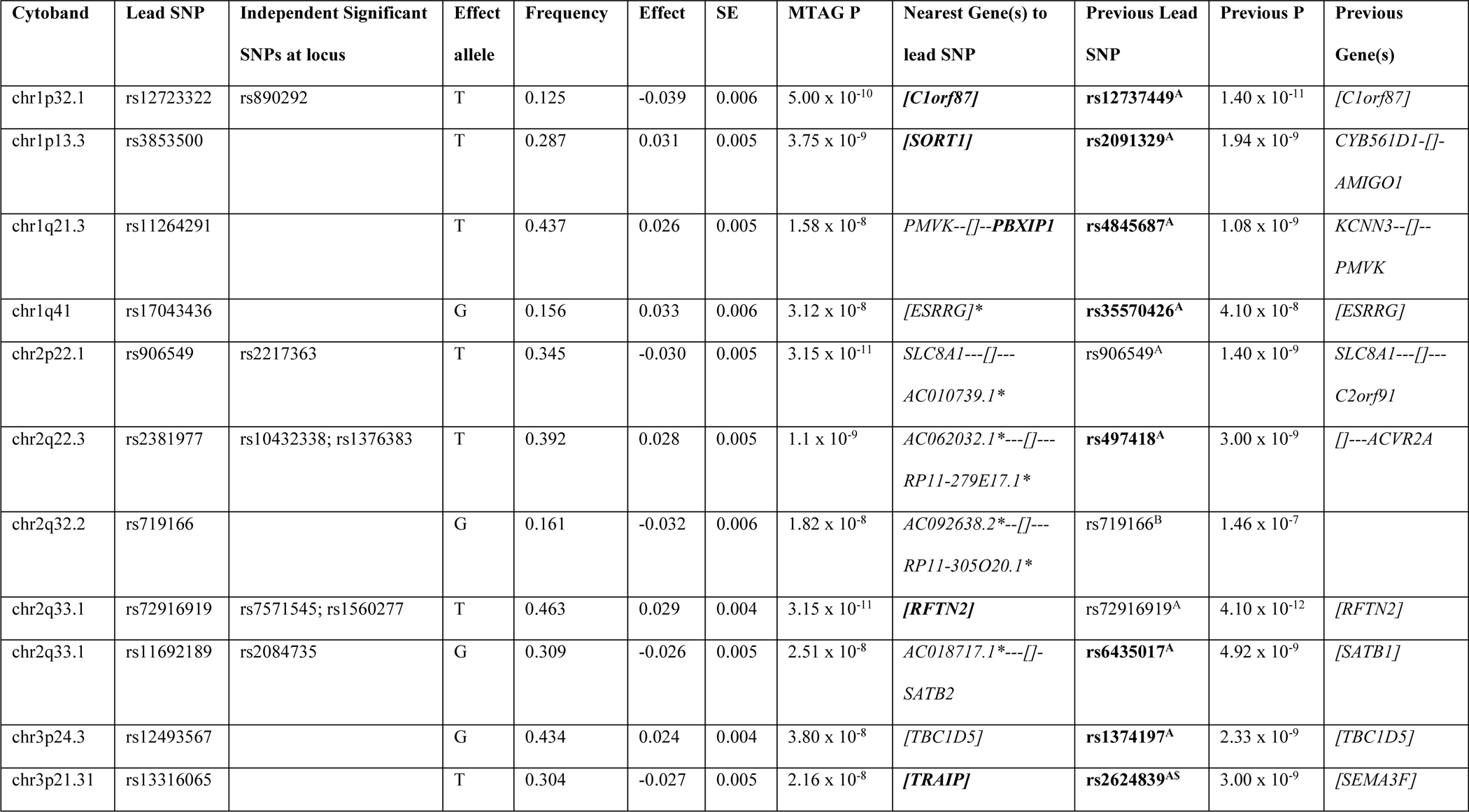
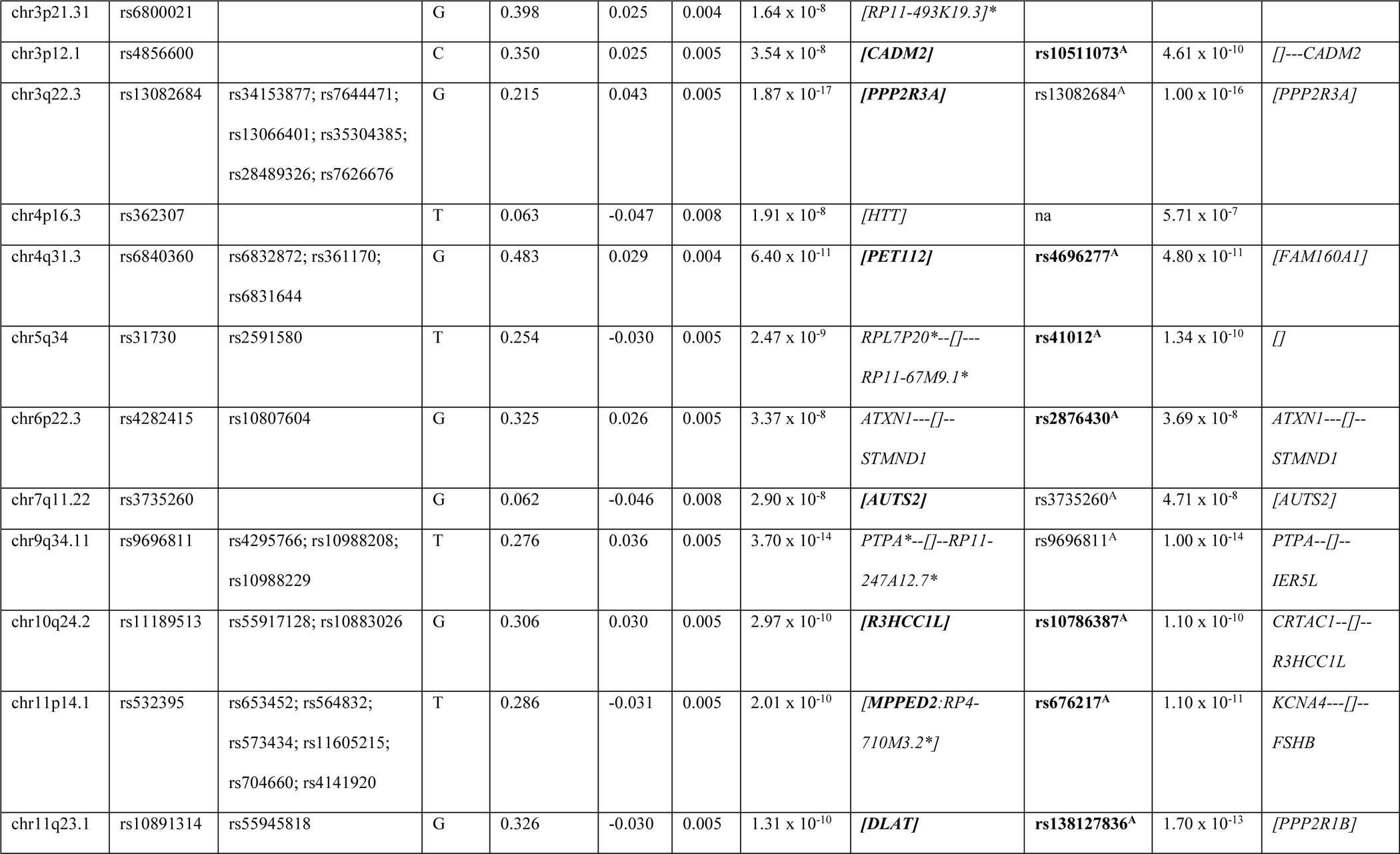
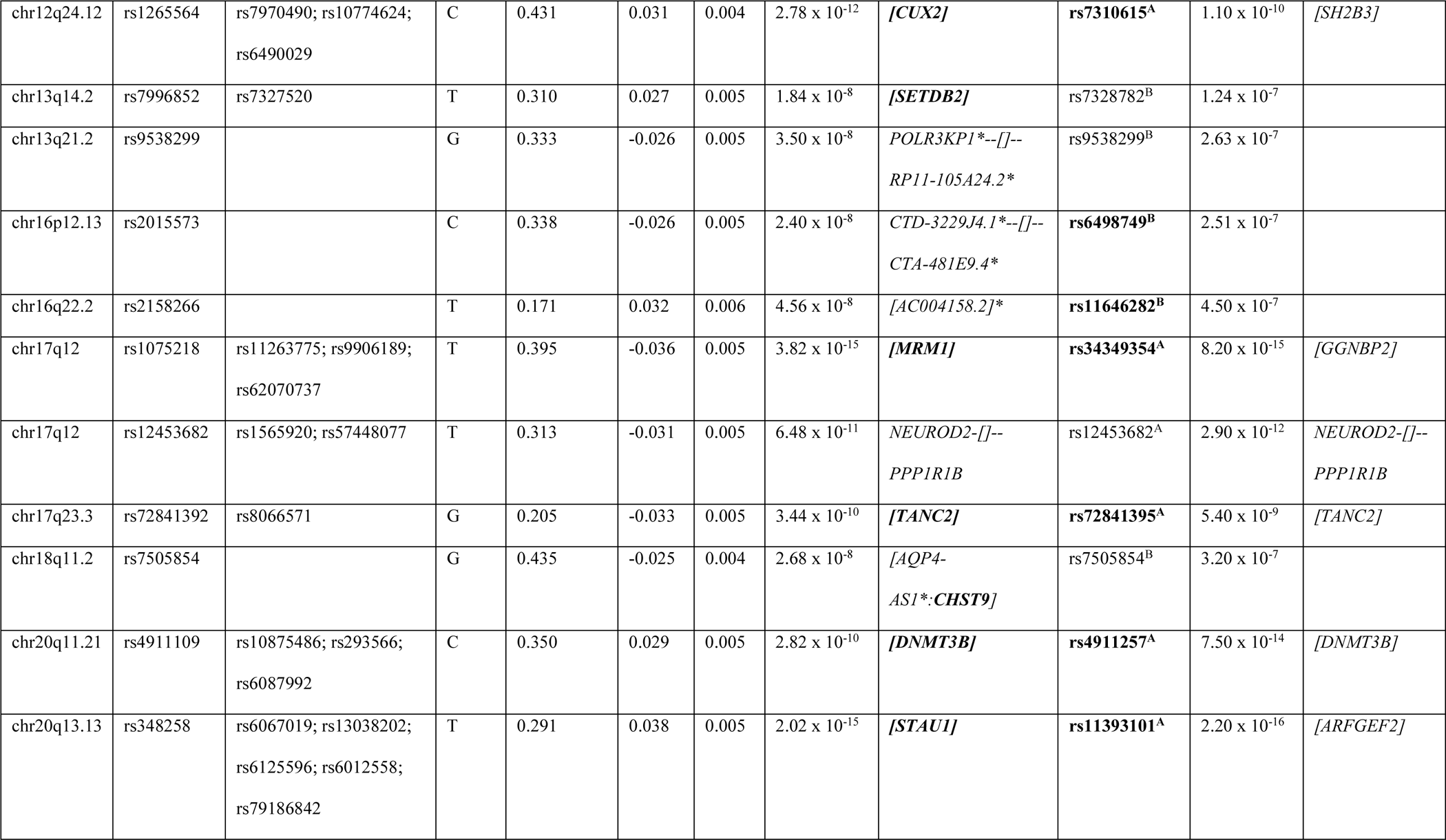
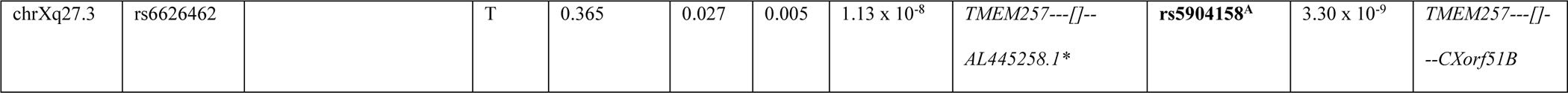
Loci associated with reading ability detected by multivariate GWAS (MTAG), with the lead SNP indicated. Genes which were significant in the gene-based test are indicated in bold, and those unavailable to test are indicated by *. Regions previously reported are indicated by A (Doust et al 2022) or B (Eising et al 2022). Where the previous lead SNP or genes have changed in the current analysis, these are indicated in bold. ^$^ indicates regions that were called as one locus in Doust et al 2022, and two in the present analysis.

The most significantly associated loci were consistent with regions previously reported in the univariate dyslexia GWAS^4^. The top locus, chr3q22.2 (rs13082684, P = 1.87×10^−17^) containing *PPP2R3A* (table 1, supplementary figure 2-14) is consistent with the top SNP of the dyslexia GWAS, although the association has become more highly significant in the MTAG analysis. Both the second and third top loci, chr20q13.13 (rs348258) (supplementary figure 2-34) and chr17q12 (rs1075218) (supplementary figure 2-30), also show more significant associations than the dyslexia source GWAS. However, for both loci, the top SNPs have changed and fall within different genes.

The lead SNP identified by the GenLang word-reading meta-GWAS^5^, rs11208009, did not reach genome-wide significance in the present multivariate study (P = 2.71×10^−6^). However, it fell within a region that reached suggestive significance (chr1:62900811-63199936) at P = 1.96×10^−7^ in which rs1168114 (LD = 0.636) was now the lead SNP. This suggestive locus overlaps completely with the original study, therefore including candidate genes *DOCK7*, *ANGPTL3*, and *USP1* (supplementary figure 3).

### Novel Regions Associated with Reading Ability

Our multivariate analysis detected seven novel regions that did not previously reach genome-wide significance in either univariate GWAS for word reading or dyslexia (table 2). One of the lead SNPs in these regions, rs362307, has been previously linked to a range of phenotypes, the most relevant of which include educational attainment^31^, cognitive ability^32^, and a “worry” phenotype key to neuroticism^33, 34^. The remaining six regions all contained lead SNPs detected by the Doust *et al* dyslexia univariate GWAS^4^ and present in the GWAS Catalogue, but at suggestive significance threshold of P ≤1×10^−5^. Interestingly, SNP rs2158266 in region chr16:72333576-72697419 containing gene *AC004158.2*, was previously associated with phoneme awareness, considered a cognitive marker of dyslexia^35^, although we note that the discovery cohort from that prior study was included in the GenLang word reading GWAS, and therefore contributed also to the current investigation.

**Table 2:** Novel regions associated with reading ability from multivariate GWAS using MTAG. Table shows associated region, lead SNP, minimum P-value, and the number of significant SNPs within the regions. The closest gene to the Lead SNP is listed, as well as other genes falling within the associated region. Relevant known associations output from FUMA are listed for both the SNP and the gene, listed as SNP: gene.

| Associated region | Lead SNP | P | Number of GWAS sig SNPs | Top SNP gene(s) | Other genes in region | Relevant known associations (SNP: gene) |
| --- | --- | --- | --- | --- | --- | --- |
| chr2:193965614-194320930 | rs719166 | 1.82 x 10<sup>-8</sup> | 11 | *AC092638.2* | *RP11-305O20.1* | Dyslexia (not GWsig) : Dyslexia (not GWsig) |
| chr4:3142660-3273010 | rs362307 | 1.91 x 10<sup>-8</sup> | 3 | *HTT* | *MSANTD1* | Educational attainment, general cognitive ability, type 2 diabetes, household income, predicted visceral adipose tissue, automobile speeding propensity, brain morphology, worrying too long after an embarrassing experience, worry/vulnerability (special factor of neuroticism), walking pace, drinks per week, years in education : as for SNPs |
| chr13:50002976-50101159 | rs7996852 | 1.84 x 10<sup>-8</sup> | 101 | *SETDB2* | *CADB39L, PHF11* | None : Dyslexia (not GWsig), mean spheric corpuscular volume, mean corpuscular volume, AFP levels |
| chr13:59485233-59706143 | rs9538299 | 3.50 x 10<sup>-8</sup> | 113 | *POLR3KP1* | *RP11-105A24.2* | Dyslexia (not GWsig) : Age at first sexual intercourse, externalizing behaviour, chronotype, intelligence, dyslexia (not GWsig) |
| chr16:17886842-18050926 | rs2015573 | 2.40 x 10<sup>-8</sup> | 55 | *CTD-3229J4.1* | *CTA-481E9.4* | None : Dyslexia (not GWsig), self-reported math ability, smoking initiation, cognitive empathy, disruptive behaviour, externalizing behaviour |
| chr16:72333576-72697419 | rs2158266 | 4.56 x 10<sup>-8</sup> | 150 | *AC004158.2* | *LINC01572, RNU6-385P, AC092289.1* | None : Dyslexia (not Gwsig), intelligence, height, smoking initiation, risk-taking, bmi, age at first sexual intercourse, phoneme awareness, schizophrenia |
| chr18:24627620-24652744 | rs7505854 | 2.68 x 10<sup>-8</sup> | 14 | *CHST9* | *AQP4-AS1* | Dyslexia (not GWsig) : Dyslexia (not GWsig) |

### Multivariate GWAS of Dyslexia

We also examined the dyslexia output of MTAG which produced an effective sample size of 1 228 832 individuals and identified 80 independent loci that met the genome-wide significance threshold (figure 1b). Of these 80 associated loci, 41 were genome-wide significant loci in the original univariate dyslexia GWAS, and 19 were reported as suggestively significant^4^. The 7 novel loci presented in our reading ability multivariate GWAS, including lead SNP rs362307 in *HTT*, were also significantly associated in our dyslexia multivariate analysis. Finally, this analysis identified 13 novel loci, not previously associated with dyslexia. These novel regions are presented in supplementary table 2. Summary statistics for SNPs reaching suggestive significance (P ≤ 1×10^−5^) are presented in supplementary table 3. Subsequent genetic and biological analyses are focussed on the reading-ability multivariate summary statistics due to the substantial increase in power for follow-up analyses, as compared to the original univariate GWAS.

### Heritability and Genetic Correlations of Reading Ability

LDSC analysis of the reading-ability multivariate GWAS revealed a high SNP-based heritability estimate (*h*^2^_snp_) of 0.24 (SE = 0.01), an increase on the previous estimate of 0.19 (SE = 0.02) previously reported for word reading^5^. Modest evidence of population stratification was indicated by lambda genomic-control estimate of 1.28, while mean χ^2^ and genomic correction having been applied by MTAG as reflected by the intercept ≤1 (0.84) and ratio <0.

Genetic correlations were estimated between multivariate reading ability and 1438 traits including recently published summary statistics for ASD^13^ and ADHD^14^. Statistically significant genetic correlations (Bonferroni corrected threshold of P ≤ 3.48×10^−5^) were found for 305 traits. A subset of relevant significant correlations is presented in figure 2, with full results in Supplementary table 4.

**Figure 2:**
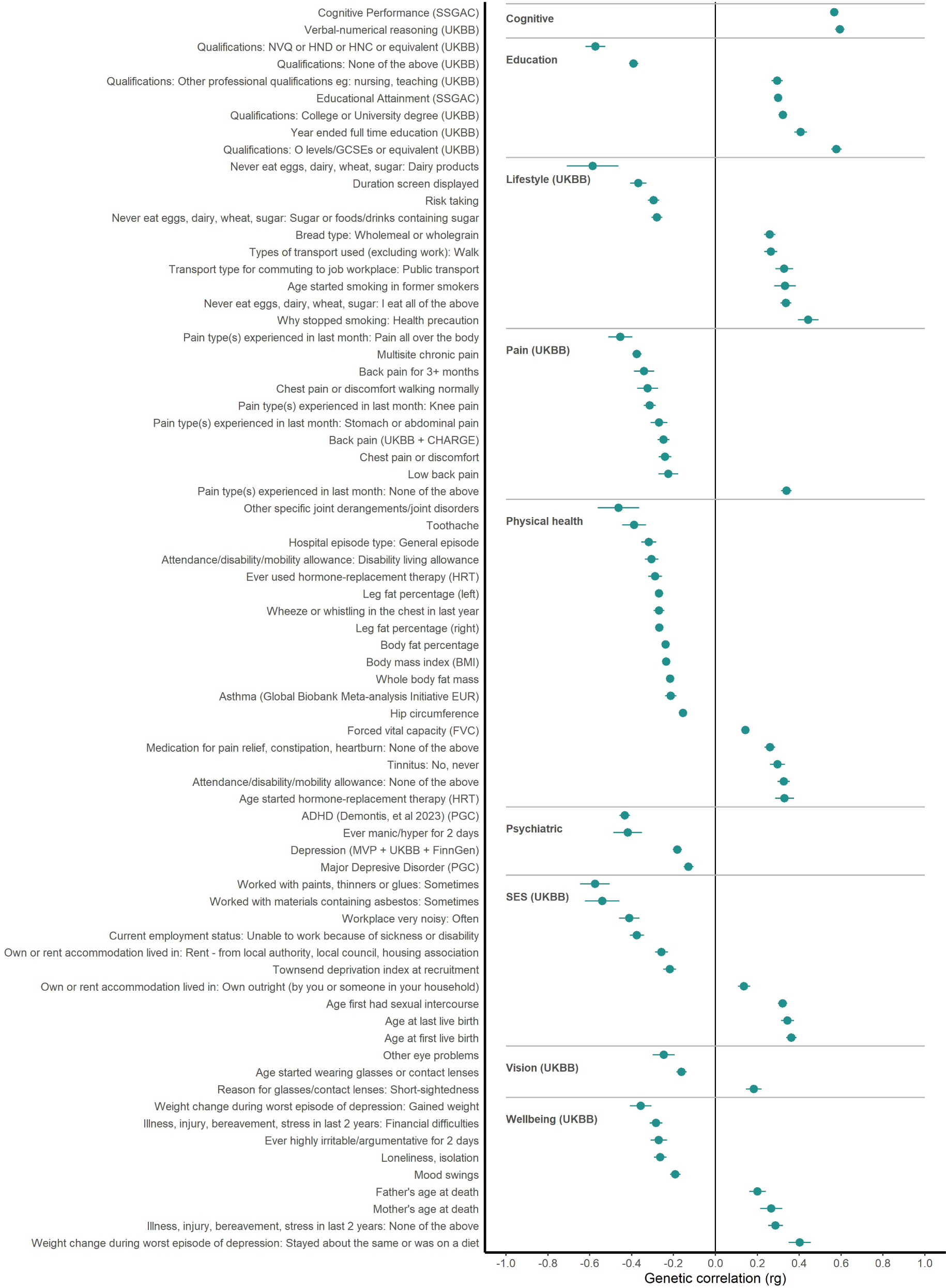
Genetic correlations of reading ability with selected relevant phenotypes. Significant (P ≤ 3.561 x 10^−5^) genetic correlations (r_g_) between multivariate analysis of reading ability and other selected phenotypes

Reading ability showed strongly positive correlations (*r*_g_) with measures of intelligence, specifically cognitive performance (0.57, SE = 0.02) (SSGAC) and verbal-numerical reasoning (UKBiobank) (0.61, SE = 0.02). Consistent with previous findings, higher reading ability was positively genetically correlated with academic achievement and education level, including educational attainment (0.30, SE = 0.02) (SSGAC), completing college or a university degree (0.32, SE = 0.02) and completing GCSEs or equivalent (0.58, SE = 0.02). Reading ability showed negative genetic correlations with achieving either vocational qualifications (NVQ or HND or HNC: −0.57, SE = 0.05; CSEs or equivalent: −0.48, SE = 0.04) or not completing any qualifications (−0.39, SE = 0.02).

In terms of neurodevelopmental traits, ADHD^14^ showed a negative correlation with reading ability (−0.43, SE = 0.03). This was weaker than that shown by the dyslexia study alone (0.53, SE = 0.12)^4^, and while not reported by Eising *et al*^5^, we calculated *r_g_* for the univariate word-reading GWAS as −0.40 (SE = 0.05). In contrast, ASD did not show a significant correlation (−0.08, SE = 0.04, P = 3.15×10^−2^) with reading ability.

Major depressive disorder showed a negative correlation (−0.13, SE = 0.03), as did measures linked to poorer wellbeing including manic episodes, risk taking and mood swings. Measures of pain and use of pain medications were also negatively correlated, including toothache (−0.39, SE = 0.06), pain all over the body (−0.45, SE = 0.06) and back pain (UKBB) (−0.24, SE = 0.03). A moderately strong positive correlation was seen between reading ability and no pain experienced in the last month (0.34, SE = 0.03). Reading ability showed negative genetic correlations with measures of lower socio-economic status and less desirable workplace conditions, and positive correlations with higher socioeconomic and health measures.

As the two constituent univariate summary statistics were derived from GWAS efforts with different sample sizes (N = 27 180 versus N_cases_ = 51 800/ N_controls_ = 1 087 070) and type of measure (quantitatively assessed continuous measure versus self-reported binary), we investigated whether the multivariate GWAS for reading ability was overpowered by signal from the dyslexia study. To do this we estimated genetic correlations between multivariate reading ability and univariate dyslexia (*r*_g_ = −0.98, SE = 0.00, Z = −334.76, P = 0.00), and between multivariate reading-ability and univariate GenLang word reading (*r*_g_ = 0.84, SE = 0.03, Z = 27.63, P = 4.66 x 10^−168^). Whereas these correlations suggested a bias from the dyslexia GWAS, there was clear evidence that the multivariate reading-ability results were indeed capturing the genes associated with quantitative reading ability with the genetic correlation almost reaching 0.90.

### Gene and Gene-set Associations

Gene-based analysis of the multivariate reading ability summary statistics identified 103 genes, meeting the Bonferroni-derived α level for 18,842 tests (P < 2.65×10^−6^). Most of these genes associated with reading ability (N = 79) were also present in associated regions detected by GWAS, while 24 fell outside of associated regions from the SNP-based screen (supplementary table 5). The overall number of associated genes (N = 103) was lower than the 173 detected by Doust *et al* (2022) in the gene-based testing of the original dyslexia study^4^. Fourteen genes were statistically associated in the present study but not in Doust *et al* (supplementary table 5). Only three of these genes fall within regions that met genome-wide significance in the present multivariate reading ability GWAS.

Eising *et al* (2022) did not report a gene-based association analysis. However, when we perform this using FUMA, only one gene meets the Bonferroni significance threshold (*AC079354.1*, P = 7.4×10^−7^), with candidate genes *DOCK7* and *USP1* showing the next strongest associations just above the Bonferroni threshold^5^.

MAGMA gene-set analysis detected enrichment of two biological pathways from 9 113 curated gene sets and gene ontology (GO) terms, tested with a Bonferroni-derived threshold (P < 5.49×10^−6^). The Gene-Set Enrichment Analysis term for Verhaak glioblastoma proneural (genes correlated with proneural type of glioblastoma multiforme tumours) containing 170 genes showed significant association (Beta = 0.35, SE = 0.08, P = 2.08×10^−6^). Similarly, the GO term for cellular compartment (GOCC) synapse containing 1 390 genes showed significant association (Beta = 0.12, SE = 0.03, P = 2.71×10^−6^). See supplementary table 6 for the full gene-set enrichment analysis.

### Variant Mapping and Functional Annotation

Candidate SNPs (N = 7 548) found in LD (*R*^2^ ≤ 0.6) with one of the independent significant SNPs, including tagged SNPs from the 1000 Genomes reference panel, underwent variant annotation using the Variant Effect Predictor (VEP) online tool (http://grch37.ensembl.org/). A total of 14 985 individual annotations were identified for candidate SNPs, due to multiple allelic variants per SNP. Intronic variants were most common (58%) with coding variants making up 0.68% (supplementary figure 4a). Of the coding variants, 51% were missense and 8% were stop-gains (supplementary figure 4b).

Six variants predicted as damaging by SIFT and PolyPhen and with CADD scores > 25, an indication for possible deleterious effects of the variants, were found: rs11142 (chr1:109897103) in *SORT1*, rs1983864 (chr10:100017453, tag SNP) in *LOXL4*, rs1064213 (chr2:198950240, tag SNP) in *PLCL1*, rs1130146 (chr 20:47859217, tag SNP) in *DDX27,* and rs10891314 (chr11:111916647) in *DLAT* with two allelic variants, G/A and G/T (supplementary table 7). Two of the variants with CADD scores > 25 (but not annotated by SIFT or PolyPhen) were predicted to result in stop gain changes: rs3764090 (chr13:50008301) in *AL136218.1* and rs2424922 (chr20:31386449, T/A) in *DNMT3B*.

At the gene level, 591 genes were contained within genome-wide significant regions (supplementary table 8). Sensitivity to loss-of-function was annotated with probability of loss-intolerance scores (pLI) and sensitivity to non-coding variation in regulatory sequences was annotated with non-coding residual intolerance scores (ncRVIS). One-hundred and twenty-five genes (21.2%) were predicted as loss-of-function intolerant by pLI ≥0.9, and seven (1.2%) were predicted as less tolerant to non-coding variation by ncRVIS ≥2.0. Three genes (*SIK2*, *PTPN14* and *XYLT1*) were predicted as intolerant by both metrics (pLI ≥0.9 and ncRVIS ≥2.0).

Ninety-seven genes (N = 97/591) located within associated regions showed evidence of association with expression QTLs (eQTL) in brain tissue (P ≤ 5.94×10^−5^) (supplementary table 8). The strongest eQTL associations were for *DHRS11* (P = 6.36×10^−77^), *CYB561* (P = 1.41×10^−69^) and *SETDB2* (P = 7.10×10^−52^).

### Functional Enrichment using Partitioned Heritability and Gene Property Analysis

To examine the tissue-specific expression profiles of genes implicated in reading ability, we used MAGMA gene property analysis within FUMA. Using RNA-seq data from the Genotype-Tissue Expression (GTEx) project, we found significant enrichments of genes associated with reading ability in brain tissue: 11 brain regions tested showed significantly higher expression levels of reading ability associated genes, particularly the cerebellum, cerebellar hemisphere and frontal cortex (supplementary figure 5, supplementary table 9) at a Bonferroni threshold of P < 4.03×10^−4^ corrected for 124 tests (GTEx and BrainSpan combined). As MAGMA analysis corrects for the average expression level in the dataset, a significant association indicates that genes associated with reading ability have a higher expression in that tissue relative to the average expression within the dataset.

Next, we tested for enrichment within the BrainSpan data set, consisting of RNA-seq from 11 developmental stages (supplementary figure 6, supplementary table 10) and 29 ages (supplementary figure 7, supplementary table 11) of human brains. No associations met Bonferroni correction for 124 tests (P < 4.03×10^−4^).

To further investigate enrichment of gene expression within the developing human brain, we tested for associations with specific cell types in single cell RNA-seq (scRNA) data using MAGMA within FUMA. Embryonic ventral midbrain from 6-11 post conception weeks (pcw) embryos (GSE76381) revealed enrichment in three cell types at Bonferroni corrected threshold of P < 6.58×10^−4^ for 76 tests (figure 3a, supplementary table 12). These were GABAergic neurons (Gaba, P = 1.07×10^−5^), neuroblast GABAergic neurons (NbGaba, P = 3.79×10^−4^), and red nucleus neurons (RN, P = 2.03×10^−4^). Embryonic prefrontal cortex scRNA-seq data from pcw 4-26 (GSE104276) showed significant enrichment in GABAergic neurons at 26 pcw (P = 5.00×10^−7^), neurons at 26 pcw (P = 2.85×10^−4^), and neurons at 16 pcw (P = 5.59×10^−5^), meeting the Bonferroni corrected threshold (76 tests, P <6.58×10^−4^) (figure 3b, supplementary table 13). Finally, scRNA-seq data from foetal and adult human cortex (GSE67835) showed significant expression in adult cortex neurons (P = 1.10×10^−5^) (76 tests, P <6.58×10^−4^) (figure 3c, supplementary table 14).

**Figure 3:**
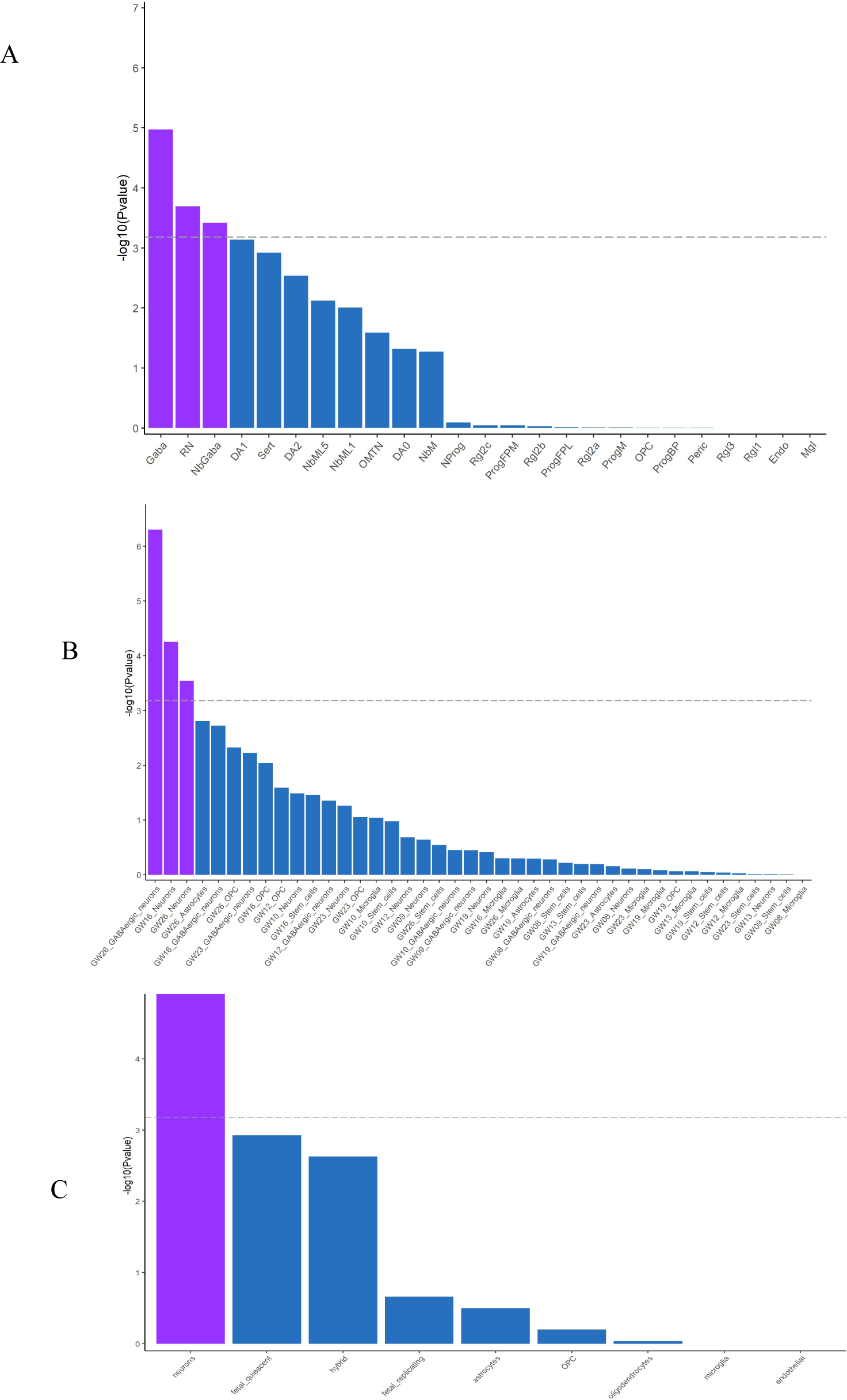
MAGMA gene property analyses of reading ability associated genes with single cell gene expression data from a) embryonic ventral midbrain from 6-11 post gestational weeks (pgw), b) embryonic prefrontal cortex from 4 – 26 pgw, c) human fetal and adult cortex. Neuronal cell lineages in a) are: DA0-1 – dopaminergic neurons, Endo – endothelial cells, Gaba – GABAergic neurons, Mgl – microglia, NbGaba - neuroblast GABAergic, NbM – medial neuroblast, NbML1-5 – mediolateral neuroblasts, NProg – neuronal progenitor, OMTN - oculomotor and trochlear nucleus, OPC – oligodendrocyte precursor cells, Peric – pericytes, ProgBP – progenitor basal plate, ProgFPL – progenitor medial floorplate, ProgM – progenitor midline, Rgl1-3 – radial glia-like cells, RN – red nucleus, Sert – serotonergic

Heritability partitioning by LDSC identified statistically significant enrichment of variants associated with reading ability in chromatin signatures annotated in foetal and adult brain tissues obtained from the Roadmap Epigenomics project and Enhancing GTEx project (ENTEx) (figure 4, supplementary table 15). Out of 489 chromatin signatures tested, twenty-nine annotations were significantly enriched (Bonferroni corrected threshold P <1.02×10^−4^), from foetal brain (N = 6), adult brain (N = 21) and primary neuronal cultured cell lines (N = 2), across a range of chromatin signatures of (active) enhancers and promoters and actively transcribed regions.

**Figure 4:**
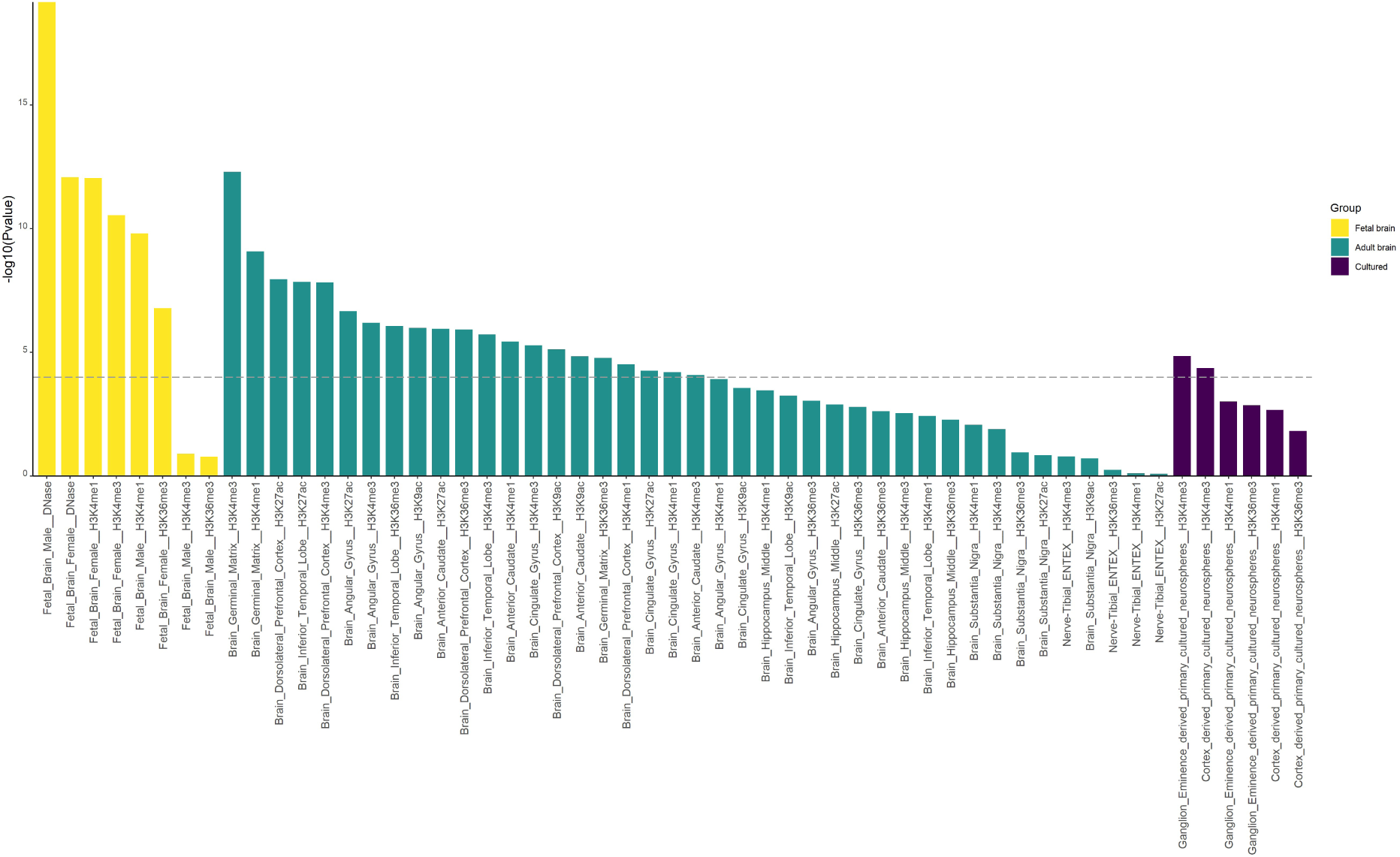
Partitioned heritability enrichment analysis of chromatin signatures. SNP-based heritability of the multivariate reading-ability GWAS is significantly enriched in brain enhancers, promoters and transcribed regions. 489 annotation of tissue-specific chromatin signatures were used to analyse the GWAS results with LDSC heritability partitioning. Only brain-related annotations are shown. P values are plotted on the y axis as −log_10_.

### Polygenic Scores Prediction in NCDS

Polygenic scores from the multivariate GWAS of reading ability were computed in an independent cohort across five developmental stages, plus an overall composite measure. The reading-ability PGS explained between 2.29 and 3.50% of variance in reading ability in the NCDS cohort. Predictions for composite measures of reading proficiency were 2.78% at age 7 years (N = 5 712), 2.61% at age 11 years (N = 5 528), and 2.29% at age 16 years (N = 4 809). Binary measures of struggling with reading scored highest at 3.5% at age 23 (N_cases_ = 167, N_controls_ = 5 288) and 2.55% at age 33 (N_cases_ = 203, N_controls_ = 5 497). The prediction for the overall composite measure of reading proficiency across all ages was 3.32% (N = 3 089) (supplementary figures 8-13; supplementary table 16).

### Polygenic Selection Analysis

The polygenic selection analysis examined if there was evidence of selection on alleles associated with reading ability seen through the past 15 000 years of human history. Essentially, this looks for differences in allele frequencies in variants associated with a trait, between the ancient ancestral population(s) and the present day. Such shifts in frequency indicate that selection acted to change the allele frequency of that variant in response to environmental pressures. From the 86 significant independent variants within 35 loci associated with reading ability in the multivariate GWAS, 42 were retained after clumping and present at high quality in the previously generated imputed ancestral data set^18, 19, 28^. Overall analyses of these SNPs identified no evidence of directional selection acting on reading ability over the past 15 000 years (P = 0.89) (figure 4). Of the 42 SNPs submitted for analysis, eight SNPs individually showed statistically significant evidence of directional selection, after accounting for the number of tests (Bonferroni threshold of P < 1.12×10^−3^ for 42 tests) (supplementary table 17).

rs10774624 in gene *LINC02356* showed the most pronounced increase in allele frequency over time and showed a positive effect on reading ability. The variant is also associated with a wide range of traits including arterial disease^36^ and rheumatoid arthritis^37^. Two further SNPs associated with positive effects on reading ability were rs1317140 in gene *TRAIP* (proxy to rs13316065), which is also associated with body mass index^38^ and urate measurement^39^, and rs7204631 with no reported association. However, both these SNPs showed a decrease in allele frequency over time.

The strongest signal in alleles associated with poorer reading ability was rs17687323 on chromosome 17 with no prior associations with other traits and showed a reduction in allele frequency over time. The remaining four SNPs also associated with poorer reading performance, were at increased frequency in modern populations; rs72916919 (no reported associations with other traits), rs9879531 in gene *NCK1* (linked to blood urea nitrogen measurement^40^), rs787995 (no association), and rs9880211 in *STAG1* (previously associated with body height^41^). It is plausible that these eight variants individually showed modest directional selection because of their contributions to traits other than reading ability, given that our overall analyses showed no evidence of selection for the latter through the past 15 000 years.

**Figure 5:**
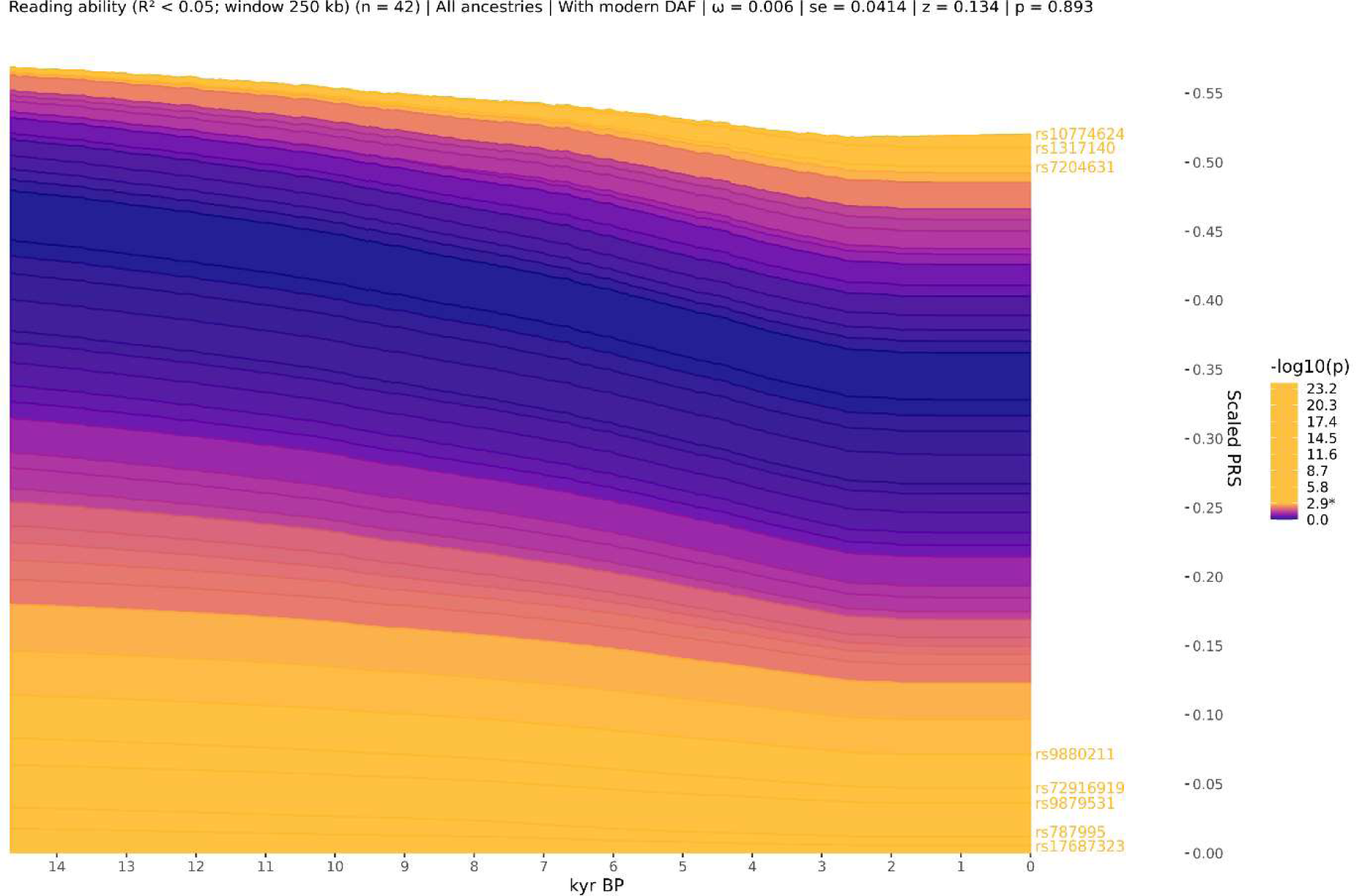
No evidence for directional selection of reading ability associated SNPs. Stacked line plot of the ancient ancestry PALM analysis, showing the contribution of SNPs to reading ability over time. SNPs are shown as stacked lines, the width of each line being proportional to the population frequency of the positive effect allele, weighted by its effect size. When a line widens over time the positive effect allele has increased in frequency, and vice versa. SNPs are sorted by the magnitude and direction of selection, with positively selected SNPs at the top, negatively selected SNPs at the bottom, and neutral SNPs in the middle. SNPs are coloured by their corresponding P-value in a single locus selection test. The asterisk on the scale bar marks the Bonferroni corrected significance threshold, and nominally significant SNPs are shown in yellow and labelled by their rsIDs. The Y-axis shows the scaled average polygenic score (PGS) in the population, ranging from 0 to 1, with 1 corresponding to the maximum possible average PGS (i.e. when all individuals in the population are homozygous for all positive effect alleles) and the X-axis shows time in units of thousands of years before present (kyr BP).

## Discussion

We performed a multivariate GWAS using MTAG on reading ability using summary statistics from the two largest reading-related cohorts to increase the effective population size to 102 082, which is substantially larger than previous studies. We detected 35 independent loci associated with reading ability, including 7 novel loci that were not significantly associated in either of the original univariate GWAS studies. The majority of loci (N = 26) were previously reported in the univariate dyslexia GWAS^4^. The most significantly associated independent SNPs in the multivariate GWAS; rs13082684 (*PPP2R3A*), rs348258 (*STAU1*) and rs1075218 (*MRM1*), were consistent with loci reported in the dyslexia GWAS^4^.

The most significantly associated SNP identified by the GenLang word-reading meta-GWAS^5^, rs11208009, did not reach genome-wide significance in the present multivariate study (P = 2.71×10^−6^). It fell within a region that reached suggestive significance (chr1:62900811-63199936, P = 1.96×10^−7^), fully overlapping with the previously reported region.

We also used the multivariate approach to identify novel loci for dyslexia, increasing the effective population size to 1 228 832. We detected 80 loci using the multivariate approach including 13 novel loci, boosting 19 regions that were previously suggestive, and seven that were novel and present in both of our multivariate analyses. However, we focussed the follow-up analyses on the reading ability output, and not on dyslexia output, because the vast increase in effective sample size compared to the respective univariate analysis, providing substantially more power than in the original GenLang study.

Annotation of associated variants indicated only six genes (seven variants) that contained potentially deleterious coding variants associated with reading ability, as predicted damaging by SIFT, PolyPhen and CADD. At the gene level, of the 591 genes contained within the associated regions, only 125 (21.2%) were predicted as loss-of-function intolerant, and seven (1.2%) were predicted as less tolerant to non-coding variation. Three genes (*SIK2*, *PTPN14* and *XYLT1*) were predicted as intolerant by both metrics. *SIK2* was previously reported in the prior dyslexia GWAS^4^, where as *PTPN14* and *XYLT1* are novel. eQTL associations revealed a large number of genes (N = 97) with evidence of expression associations in brain tissue, similar to those identified in the prior dyslexia GWAS, the strongest associations were identified in genes *DHRS11* and *CYB561*.

We identified seven novel loci that were not previously significantly associated with either word reading or dyslexia in the original univariate GWASs. The novel lead SNP, rs362307 within *HTT,* has been associated with a range of cognitive traits relevant to reading ability in prior studies, including cognitive function^32^ and educational attainment^31^. Genomic structural equation modelling and genetic correlation analyses have shown that reading-related traits have substantial genetic overlaps with educational attainment and IQ^4, 5^, and therefore SNPs involved in general cognition are likely to be important across traits. The remaining six loci were identified in the univariate dyslexia study at a sub-genome-wide association threshold^4^, but are present in the GWAS Catalogue^42^. One of these regions, chr16:72333576-72697419 in gene *AC004158.2* (rs2158266) was previously reported by Gialluisi and colleagues as suggestively associated with phoneme awareness, deficits of which are considered a marker of dyslexia^35^. That study included 3 093 individuals of European ancestry from the Neurodys, CLDRC and UKdys cohorts, which also formed part of the GenLang meta-GWAS, meaning that these findings are not fully independent.

We predicted a maximum of 3.5% of trait variance in reading measures in the NCDS using the multivariate word reading polygenic scores derived from our multivariate reading ability GWAS. This is an improvement on the previous predictions using the dyslexia GWAS on measures of word reading in similar population-based cohorts where 2.9% of variance was explained in adolescents and ∼2% in adults^4^. Predictions across longitudinal measures at five ages within the NCDS ranged from 2.29-3.5% suggesting that this part of the genetic contribution of reading is stable through schooling and into adulthood. Other studies have used phenotypically related PGS to predict reading outcomes. For example, Selzam *et al* (2017)^43^ used a years-in-education PGS^44^ to predict 5% of reading variance at 14 years of age. Future studies might test whether our reading ability PGS explains incremental variance in reading variation above an educational attainment PGS, which is a broader phenotype encompassing cognitive and non-cognitive factors.

Genetic correlations between the multivariate reading-ability trait and other phenotypes showed a high degree of similarity to the profiles of genetic correlations seen for both the univariate GWAS studies of word reading and dyslexia. Key findings were consistent with the univariate GWAS, such as strong genetic correlations with educational attainment, completing a college or university degree, and a negative correlation with manual jobs. Notably, the strongest correlation for the multivariate reading-ability GWAS was verbal-numerical reasoning in the UKBiobank (0.61, SE = 0.02), which was also among the strongest correlations for Dyslexia at −0.5 (SE = 0.03)^4^.

Correlations with other neurodevelopmental traits were reasonably consistent for the univariate and multivariate reading-ability GWASs. The strongest correlation of the dyslexia GWAS was with ADHD at 0.53 (SE = 0.01)^4^, and although this correlation was lower in absolute magnitude for the multivariate reading-ability analysis (−0.43, SE = 0.03), it remains consistent with the literature linking ADHD with reading and language outcomes^9, 45^. The observation that genetic correlation with ADHD is stronger with dyslexia than with reading ability overall may indicate that dyslexia diagnosis is influenced by more than just reduced reading performance, and that these other considerations overlap with indicators of ADHD. Consistent with previous findings that there is no significant genetic correlation between autism spectrum disorder (ASD) and reading ability^4, 5^, no significant correlation was found in the multivariate reading-ability investigation carried out here.

Gene-set analysis revealed two enriched biological pathways implicated in reading ability: the gene-ontology term for cellular compartment (GOCC) neural synapses, and for Verhaak glioblastoma proneural (genes correlated with proneural type of glioblastoma multiforme tumours). Both gene-sets hint at essential neuronal mechanisms.

Our analysis of expression patterns of associated genes in the developing human brain offered further evidence for a role in early developmental processes, implicating GABAergic and red neurons in embryonic prefrontal cortex and midbrain, as well as cortex neurons in adults. These findings echo the cell type analysis reported in the GenLang paper, where an enrichment in red nucleus neurons and a trend towards enrichment in fetal GABAergic neurons was observed^5^. Price and colleagues reported evidence supporting neuronal migration/axon guidance as potential pathways using a candidate gene-set approach for known neurodevelopmental genes in a hypothesis-driven association analysis of word reading which included the GenLang meta-GWAS^46^. More recently, the same research group implicated glutamatergic (excitatory) and GABAergic (inhibitory) neurons in the adult cortex in word reading, using a subset of the GenLang cohorts (N = 5 054)^17^. The sample used in the present study overlaps with that of the prior work, which may contribute to the consistency between the two sets of results. Despite the small GWAS sample size used by Price and colleagues, their work offers support for the GABAergic inhibitory system as a future focus for connecting genetics to neuronal mechanisms.

Polygenic selection analysis found no significant selection observed from ancestral populations suggesting that the genetic influences on reading skills were not specifically selected for or against in the transition between hunter gatherer and farmers in Europeans. This finding may be considered unsurprising since reading is many thousands of years old but has only very recently become widespread and has no obvious selection pressure or effect on reproductive fitness. However, because reading processes are highly dependent on brain circuits that evolved in support of spoken language, it was still possible that we may have detected signals related to language evolution. The consistency of the PGS through the past 15k years of history in northern Europe suggests it has not been affected by any major social or societal changes that have taken place in history such as the transition to farming, although it is important to note that our PGS accounts for only a small proportion of heritable reading ability. We identified eight SNPs that showed individually significant changes through recent history; five showed evidence of negative selection over time and three showed signs of positive selection, although their direction of effect on reading-ability were mixed. Considering that reading ability is likely a neutral trait with regard to biological fitness, we speculate that the patterns observed for these eight SNPs are most likely due to selection pressures acting on pleiotropic traits.

Analysis that examines further back through evolution (30 million years ago to 50 000 years ago) was performed in both the Eising *et al*^5^ and Doust *et al*^4^ papers. Based on findings of human brain structure evolution^47^, five annotations reflecting aspects of human evolution were examined. Doust et al found no evidence of enrichment for annotations related to human evolution. Eising et al found evidence of an enrichment in archaic deserts; long regions in the human genome where there is an absence of Neanderthal admixture, suggesting these regions may be intolerant to gene flow and therefore harbouring variants essential to *Homo sapiens*. The findings suggested that these archaic desert regions could contain genetic variations that contribute more to reading and language traits in modern humans than expected by chance.

In terms of limitations of the current study, we note that the false discovery rate (FDR) was moderate at 6%, primarily because of the size imbalance between the constituent summary statistics. Genetic correlation between the multivariate reading ability phenotype was marginally stronger with dyslexia than GenLang word reading. When considered along with the MTAG modest FDR, they suggest the observed patterns of genetic associations may be influenced by the larger dyslexia GWAS.

In summary, through implementation of multivariate GWAS analysis combining work on quantitative measures with self-report of disorder, we have produced the largest genetic study of reading ability (effective N = 102 082) to date. Our findings account for 24% (*h^2^_snp_*) of the observed variability of reading ability in our datasets. We identified seven novel loci associated with quantitative reading ability and implicated early brain developmental processes in the biological underpinnings of reading.

## Data Availability

The full summary statistics for each dyslexia GWAS presented in this paper will be made available through 23andMe website (https://research.23andme.com/dataset-access/) to qualified researchers under an agreement with 23andMe that protects the privacy of the 23andMe participants. SNPs that met suggestive genome-wide significance (P<1×10-5) are made available in the supplementary tables.

## Acknowledgements

We thank the participants and employees of 23andMe, Inc, and the participants and research members of the GenLang Consortium. We are grateful to the Centre for Longitudinal Studies (CLS), UCL Social Research Institute, for the use of the NCDS data and to the UK Data Service for making them available. However, neither CLS nor the UK Data Service bear any responsibility for the analysis or interpretation of these data.

We thank Drs David Hill and Charley Xia for their assistance with the multivariate analyses and genetic correlations, and to Jonny Flint for assistance with the polygenic score analysis.

SEF and EE are supported by the Max Planck Society (Germany). EE is also supported by a Veni grant of the Dutch Research Council (NWO; VI.Veni.202.072). HSM is supported by the Biotechnology and Biological Sciences Research Council [BB/T000813/1].

The following members of the 23andMe Research Team contributed to this study:

Stella Aslibekyan, Adam Auton, Elizabeth Babalola, Robert K. Bell, Jessica Bielenberg, Jonathan Bowes, Katarzyna Bryc, Ninad S. Chaudhary, Daniella Coker, Sayantan Das, Emily DelloRusso, Sarah L. Elson, Nicholas Eriksson, Teresa Filshtein, Pierre Fontanillas, Will Freyman, Zach Fuller, Chris German, Julie M. Granka, Karl Heilbron, Alejandro Hernandez, Barry Hicks, David A. Hinds, Ethan M. Jewett, Yunxuan Jiang, Katelyn Kukar, Alan Kwong, Yanyu Liang, Keng-Han Lin, Bianca A. Llamas, Matthew H. McIntyre, Steven J. Micheletti, Meghan E. Moreno, Priyanka Nandakumar, Dominique T. Nguyen, Jared O’Connell, Aaron A. Petrakovitz, G. David Poznik, Alexandra Reynoso, Shubham Saini, Morgan Schumacher, Leah Selcer, Anjali J. Shastri, Janie F. Shelton, Jingchunzi Shi, Suyash Shringarpure, Qiaojuan Jane Su, Susana A. Tat, Vinh Tran, Joyce Y. Tung, Xin Wang, Wei Wang, Catherine H. Weldon, Peter Wilton, Corinna D. Wong.

## Conflict of Interest

PF, AA and the 23andMe Research Team are employed by and hold stock or stock options in 23andMe, Inc. All other authors declare no conflicts of interest.

Supplementary information is available at Molecular Psychiatry’s website

## Availability of Data and Materials

The univariate GWAS summary statistics for word reading are available to download from the GenLang website https://www.genlang.org/downloads.html. The full GWAS summary statistics for the 23andMe discovery data set will be made available through 23andMe to qualified researchers under an agreement with 23andMe that protects the privacy of the 23andMe participants. Please visit https://research.23andme.com/collaborate/#dataset-access/ for more information and to apply to access the data. The multivariate summary statistics for reading ability generated by this study are available through 23andMe to qualified researchers, as described.

