## Supplementary text for "Multivariate genome-wide association analysis of quantitative reading skill and dyslexia improves gene discovery"

***Supplementary Table 1****: SNPs which were most strongly associated ($P \leq 1 \times 10^{-5}$, N = 9,717) with multivariate analysis of reading ability.*

***Supplementary Table 2****: SNPs which were most strongly associated ($P \leq 1 \times 10^{-5}$, N = 18,551) with multivariate analysis of dyslexia.*

***Supplementary Table 3:*** *Novel regions associated with multivariate GWAS of dyslexia. Regions are described as either novel, also reported in the multivariate analysis of reading ability, or previously met suggestive significance in Doust et. al. 2023.*

***Supplementary Table 4:*** *Complete results for all 1438 genetic correlations tested in LDSC.*

***Supplementary Table 5:*** *Gene-based association results tested for 18,842 genes using MAGMA for multivariate reading ability.*

***Supplementary Table 6:*** *Gene-set association results for 15,486 biological pathways using MAGMA for multivariate reading ability.*

***Supplementary Table 7:*** *Variant Effect Predictions for coding SNPs. Prioritised variants annotated as damaging by both SIFT and PolyPhen2 (N = 6) are presented in bold.*

***Supplementary Table 8:*** *Gene-based annotations using FUMA showing loss-of-function predictions, and expression QTL associations.*

***Supplementary Table 9:*** *MAGMA gene-property analysis of multivariate reading ability partitioned by GTEx gene expression in individual tissues.*

***Supplementary Table 10:*** *MAGMA gene-property analysis partitioned by brain tissue gene expression across 11 developmental stages in BrainSpan.*

***Supplementary Table 11:*** *MAGMA gene-property analysis partitioned by brain tissue gene expression across 29 ages in BrainSpan.*

***Supplementary Table 12:*** *MAGMA gene-property analysis partitioned by single-cell RNA-seq brain tissue gene expression in embryonic ventral mid-brain.*

***Supplementary Table 13:*** *MAGMA gene-property analysis partitioned by single-cell RNA-seq brain tissue gene expression in embryonic pre-frontal cortex.*

***Supplementary Table 14:*** *MAGMA gene-property analysis partitioned by single-cell RNA-seq brain tissue gene expression in adult and foetal cortex grouped by neuronal cell type.*

***Supplementary Table 15:*** *Results of LDSC partitioning heritability in tissue-specific chromatin modification patterns from the Roadmap Epigenomics project and ENTEX, using the annotations and method of Finucane et al. 2015* [25].

***Supplementary Table 16:*** *Polygenic score prediction for reading-ability multivariate summary statistics across six measures of longitudinal reading ability in the National Child Development Study, 1958.*

***Supplementary Table 17:*** *Table showing polygenic selection analysis results for independent SNPs associated with reading ability in an imputed ancient panel.*
