## Supplementary figures for "Multivariate genome-wide association analysis of quantitative reading skill and dyslexia improves gene discovery"

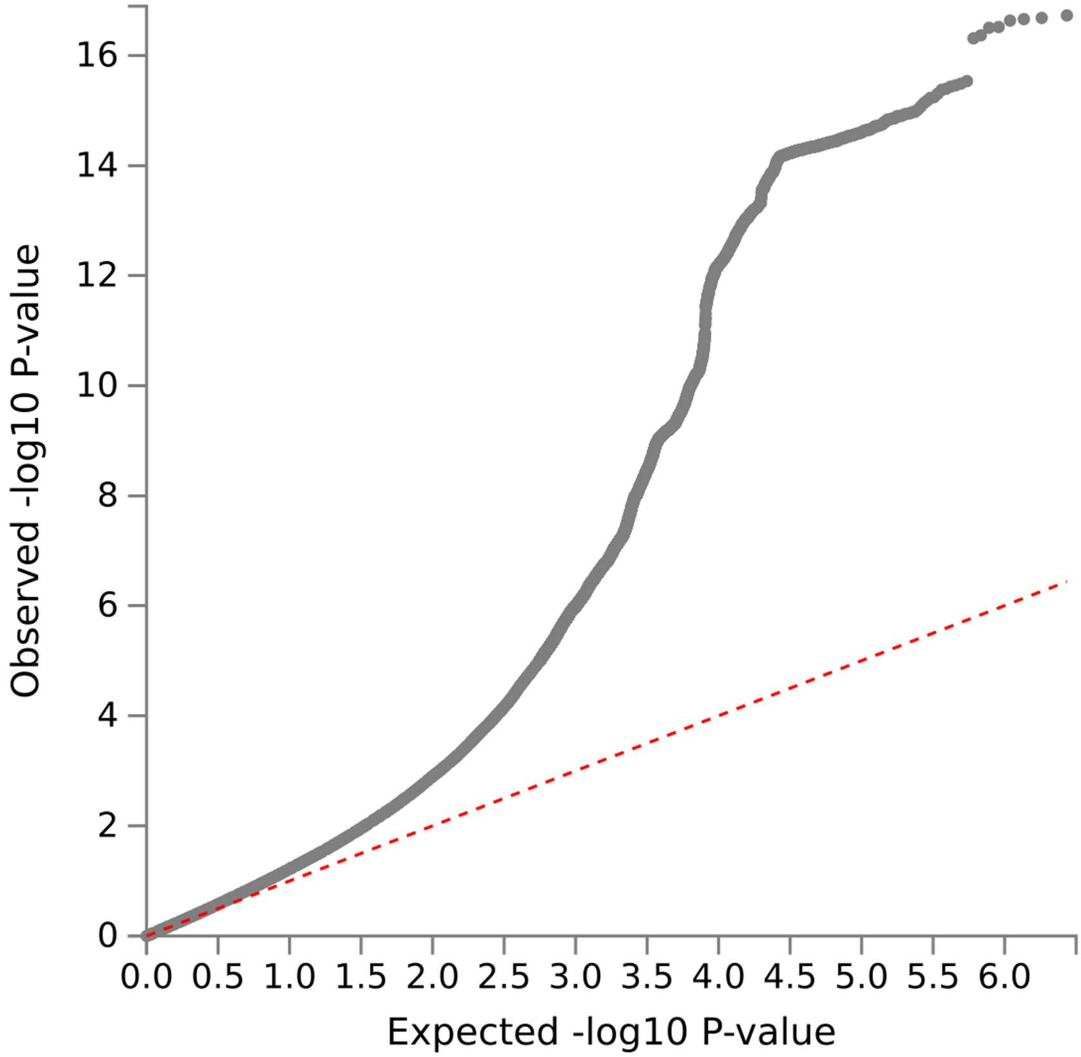


**Supplementary figure 1**: Quantile-Quantile (Q-Q) plot showing expected versus observed -log10 P-values for each variant from multivariate GWAS of reading ability, represented by grey dots. The dashed red line shows the null hypothesis. The plots indicate the absence of uncorrected population stratification.

**Supplementary figure 2-1**: LocusZoom of regions associated with reading ability, region chr1p32.1 rs12723322.

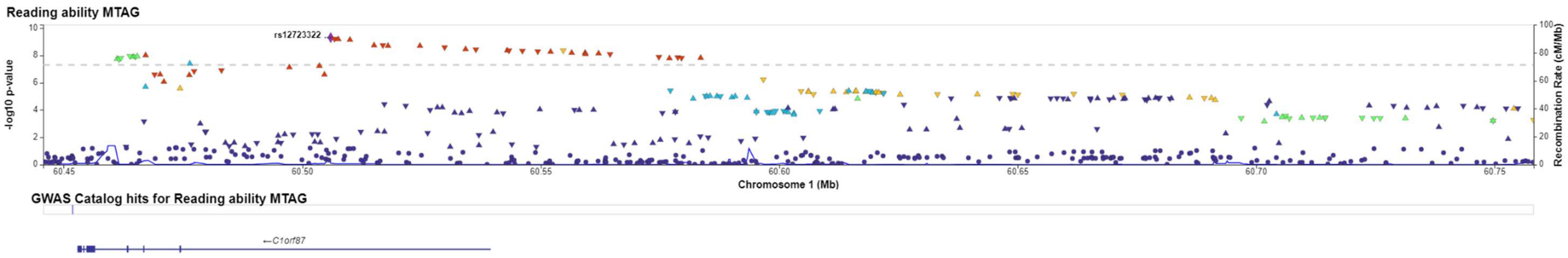


**Supplementary figure 2-2**: LocusZoom of regions associated with reading ability, region chr1p13.3 rs3853500.

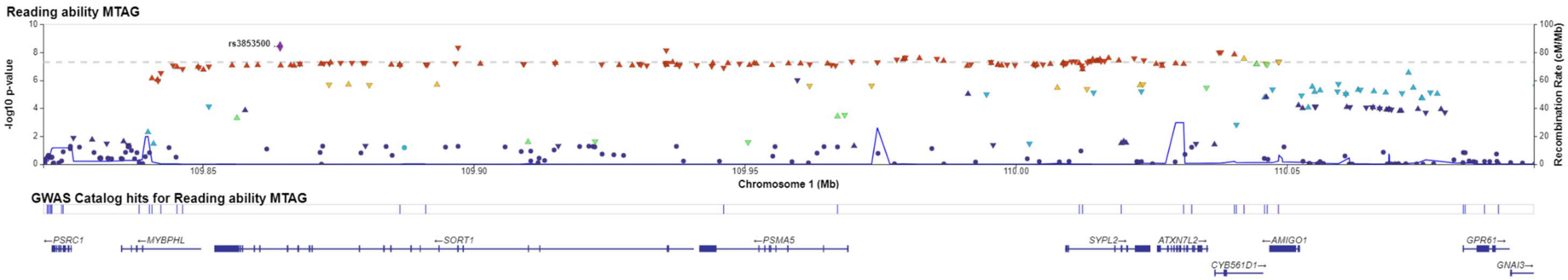

**Supplementary figure 2-3**: LocusZoom of regions associated with reading ability, region chr1p21.3 rs11264291.

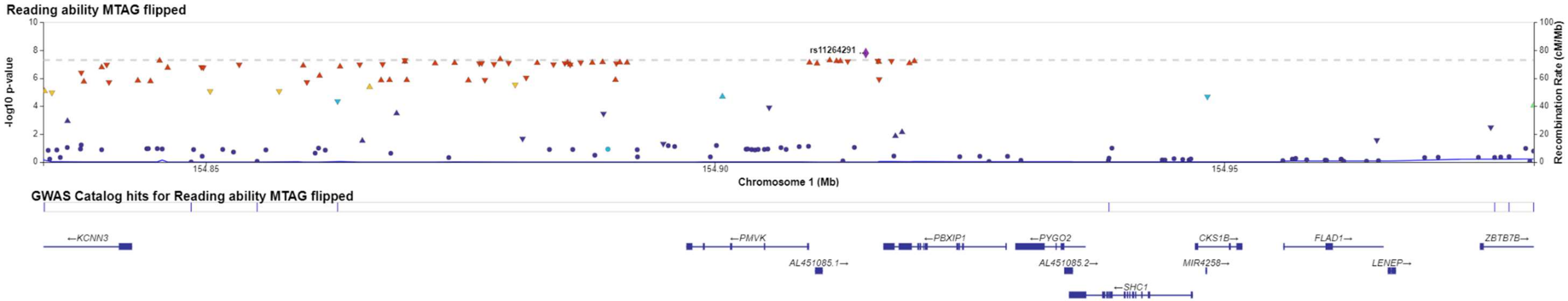


**Supplementary figure 2-4**: LocusZoom of regions associated with reading ability, region chr1q41 rs17043436.

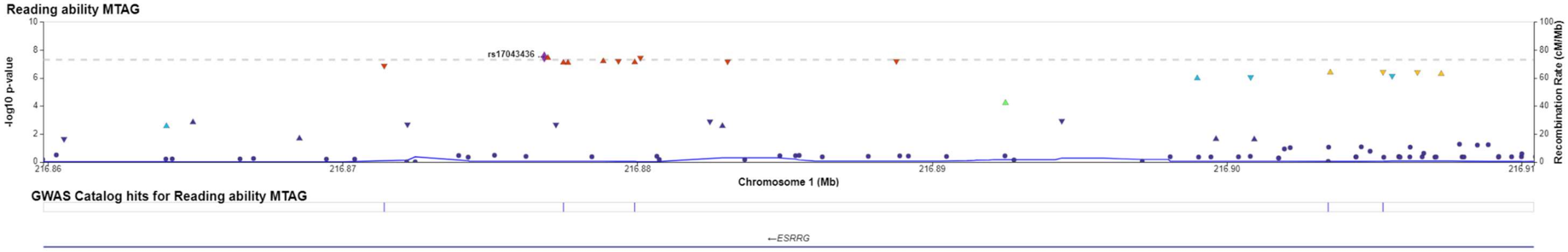

**Supplementary figure 2-5**: LocusZoom of regions associated with reading ability, region chr2p22.1 rs906549.

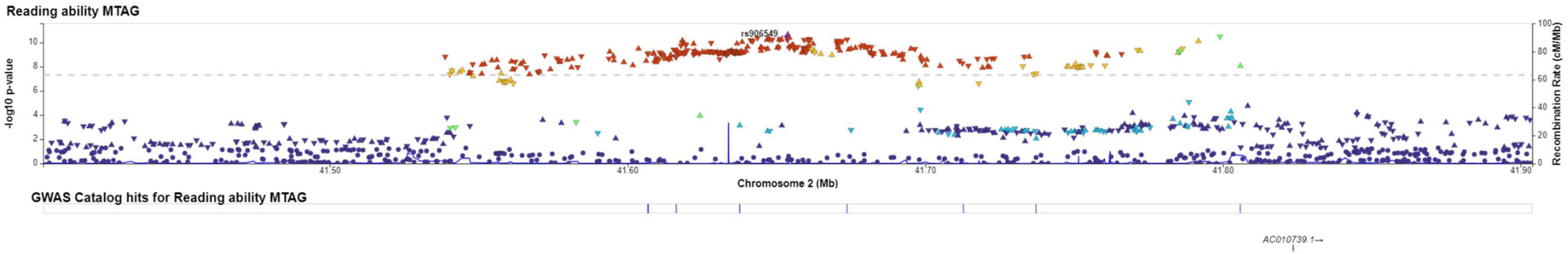


**Supplementary figure 2-6**: LocusZoom of regions associated with reading ability, region chr2q22.3 rs906549.

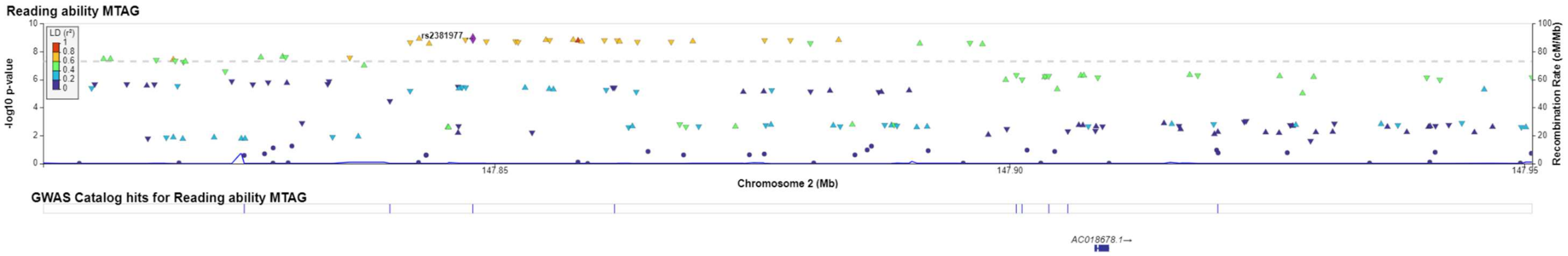

**Supplementary figure 2-7**: LocusZoom of regions associated with reading ability, region chr2q32.2 rs719166.

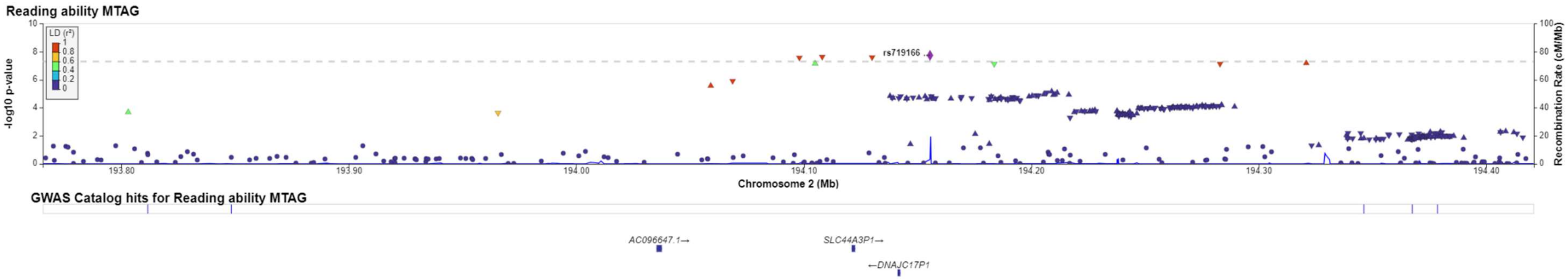


**Supplementary figure 2-8**: LocusZoom of regions associated with reading ability, region chr2q33.1 rs72916919.

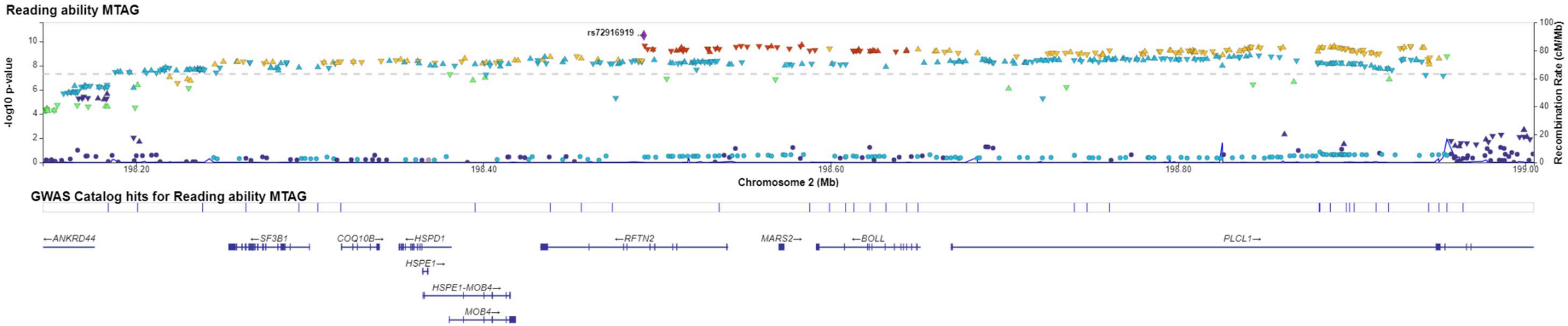

**Supplementary figure 2-9**: LocusZoom of regions associated with reading ability, region chr2q33.1 rs11692189.

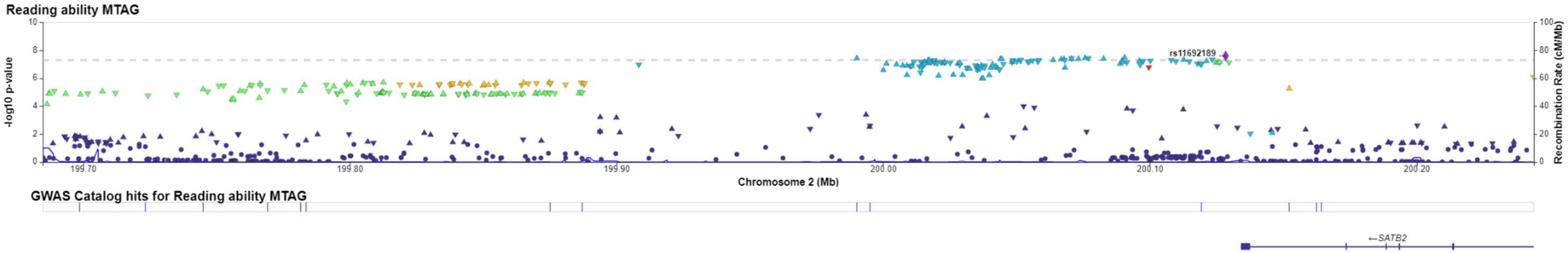


**Supplementary figure 2-10**: LocusZoom of regions associated with reading ability, region chr3p24.3 rs12493567.

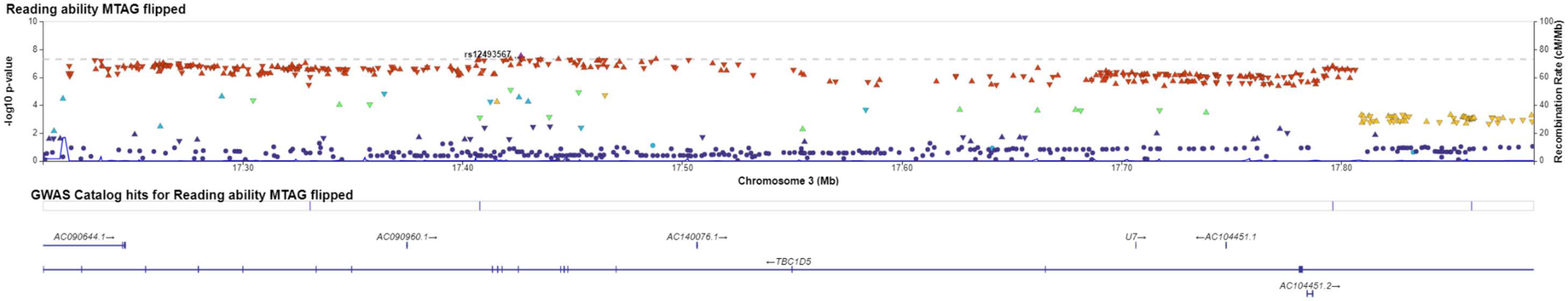

**Supplementary figure 2-11**: LocusZoom of regions associated with reading ability, region chr3p21.31 rs13316065.

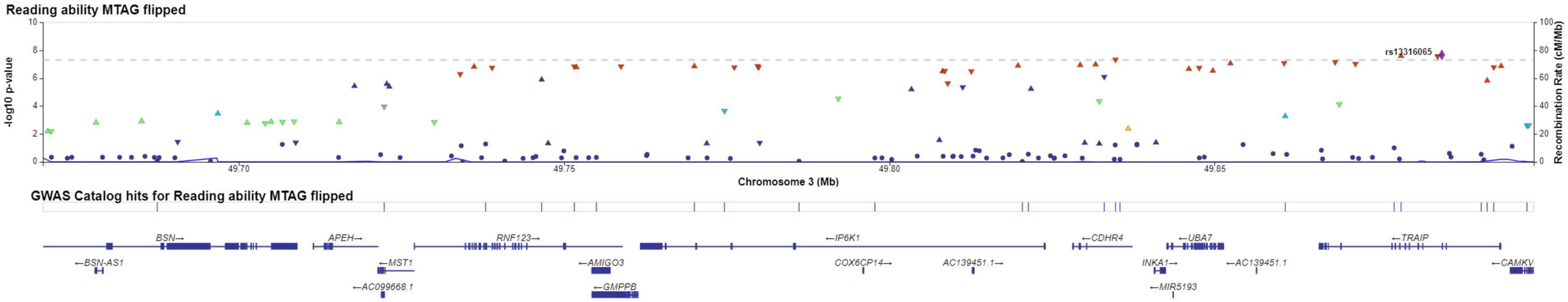


**Supplementary figure 2-12**: LocusZoom of regions associated with reading ability, region chr3p21.31 rs6800021.

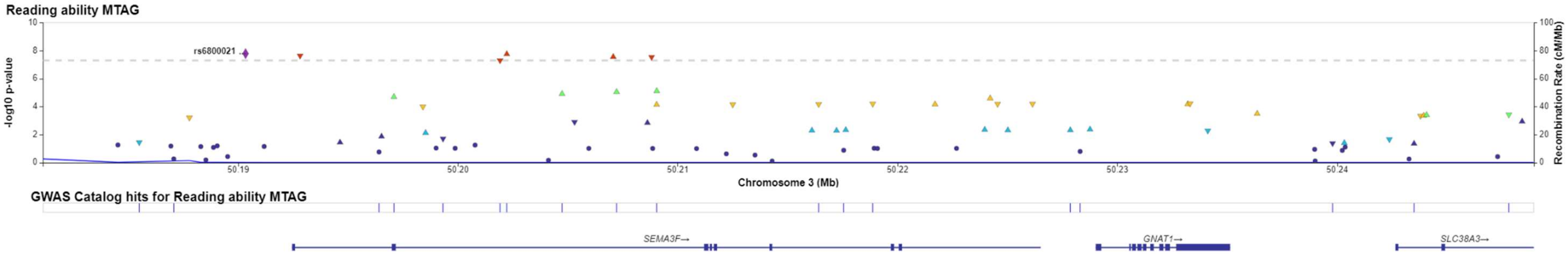

**Supplementary figure 2-13**: LocusZoom of regions associated with reading ability, region chr3p12.1 rs4856600.

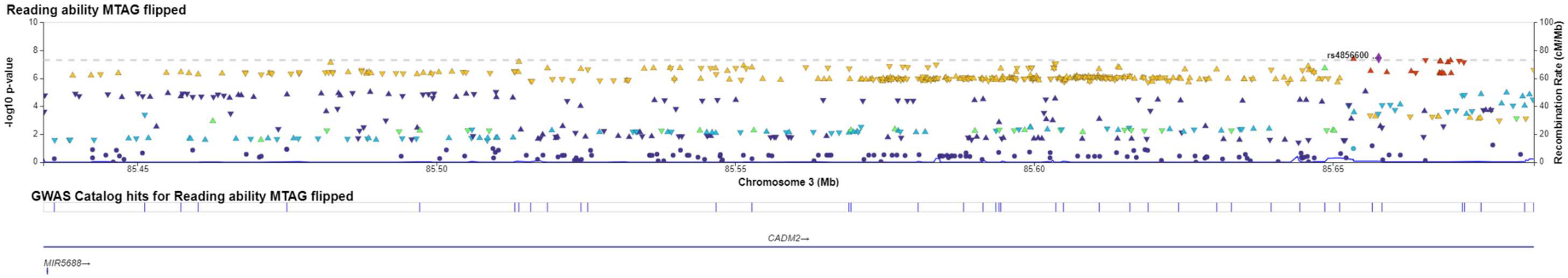


**Supplementary figure 2-14**: LocusZoom of regions associated with reading ability, region chr3q22.3 rs13082684.

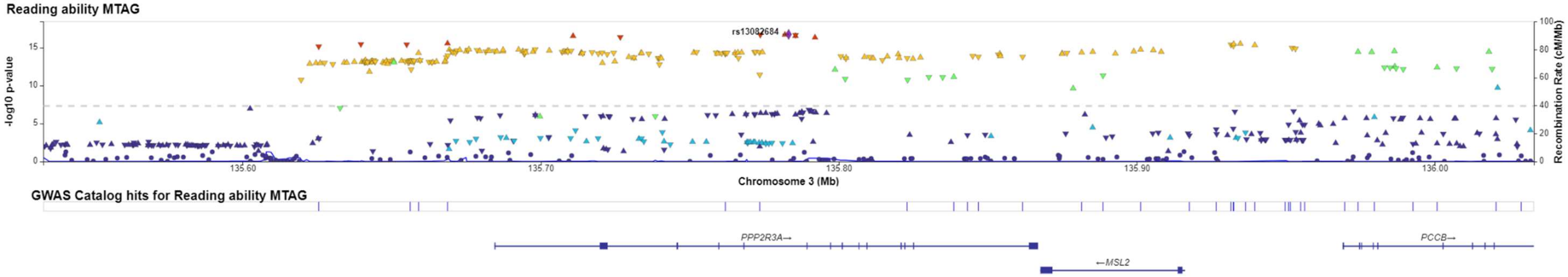

**Supplementary figure 2-15**: LocusZoom of regions associated with reading ability, region chr4p16.3 rs362307.

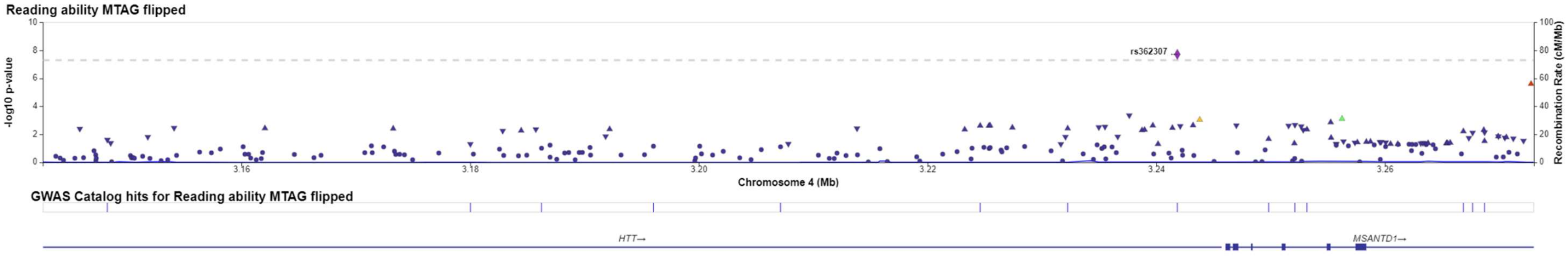


**Supplementary figure 2-16**: LocusZoom of regions associated with reading ability, region chr4q31.3 rs6840360.

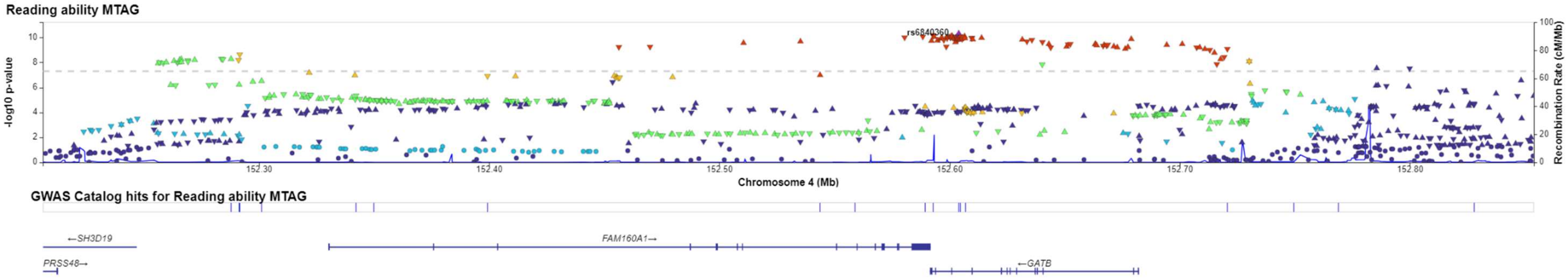

**Supplementary figure 2-17**: LocusZoom of regions associated with reading ability, region chr5q34 rs31730.

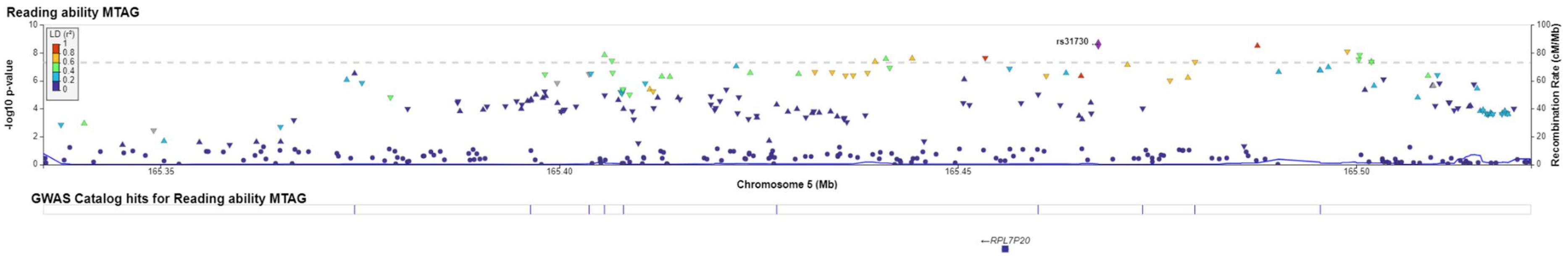


**Supplementary figure 2-18**: LocusZoom of regions associated with reading ability, region chr6p22.3 rs4282415.

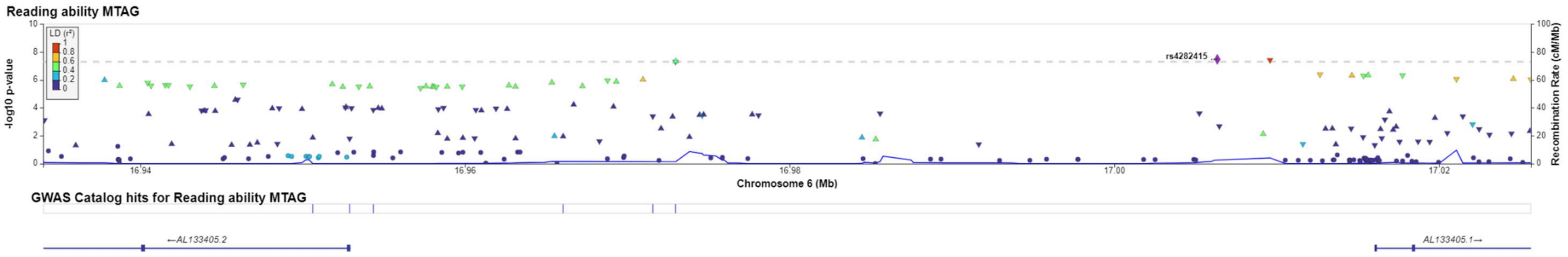

**Supplementary figure 2-19**: LocusZoom of regions associated with reading ability, region chr7q11.22 rs3735260.

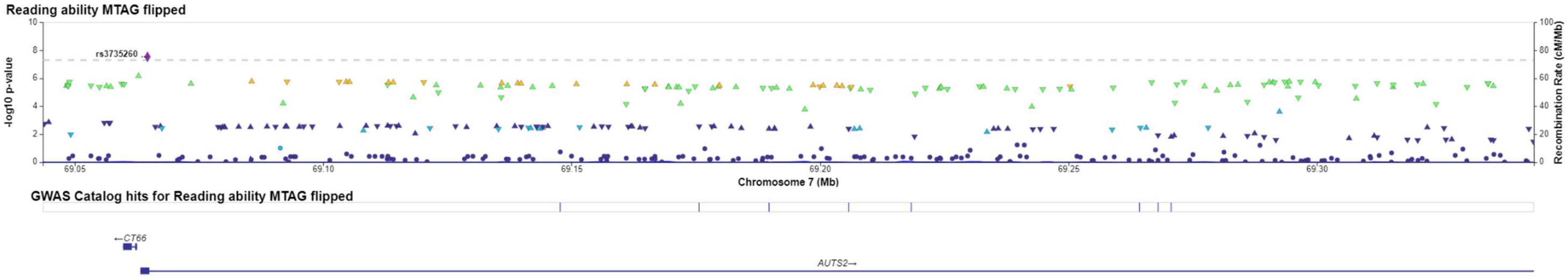


**Supplementary figure 2-20**: LocusZoom of regions associated with reading ability, region chr9q34.11 rs9696811.

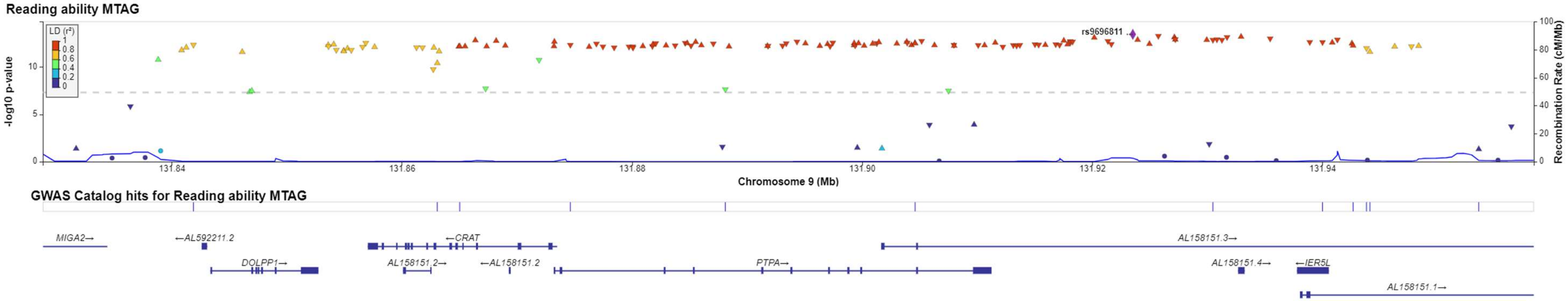

**Supplementary figure 2-21**: LocusZoom of regions associated with reading ability, region chr10q24.2 rs11189513.

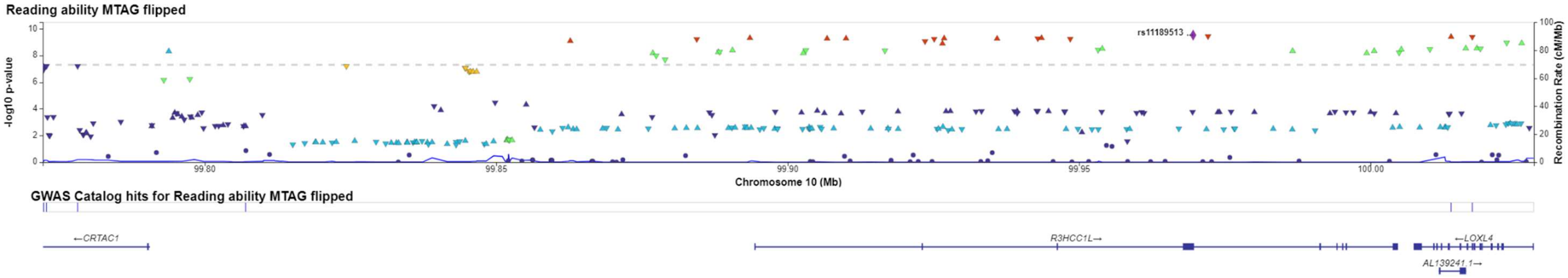


**Supplementary figure 2-22**: LocusZoom of regions associated with reading ability, region chr11p14.1 rs532395.

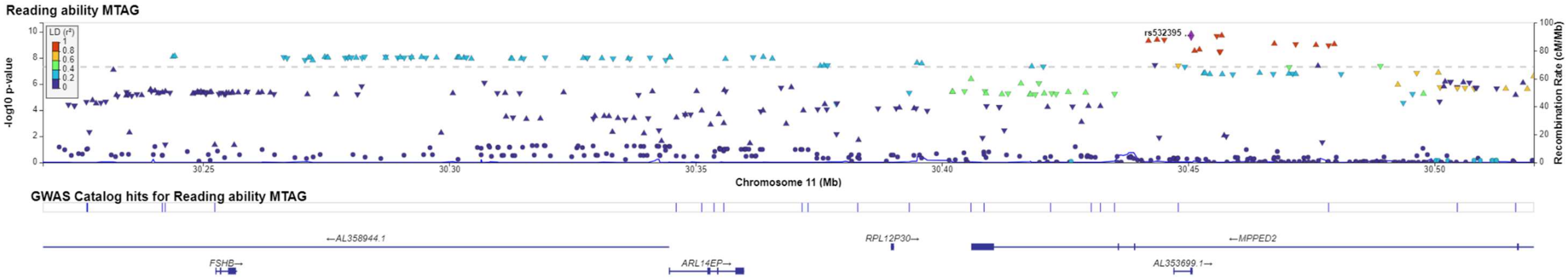

**Supplementary figure 2-23**: LocusZoom of regions associated with reading ability, region chr11q23.1 rs10891314.

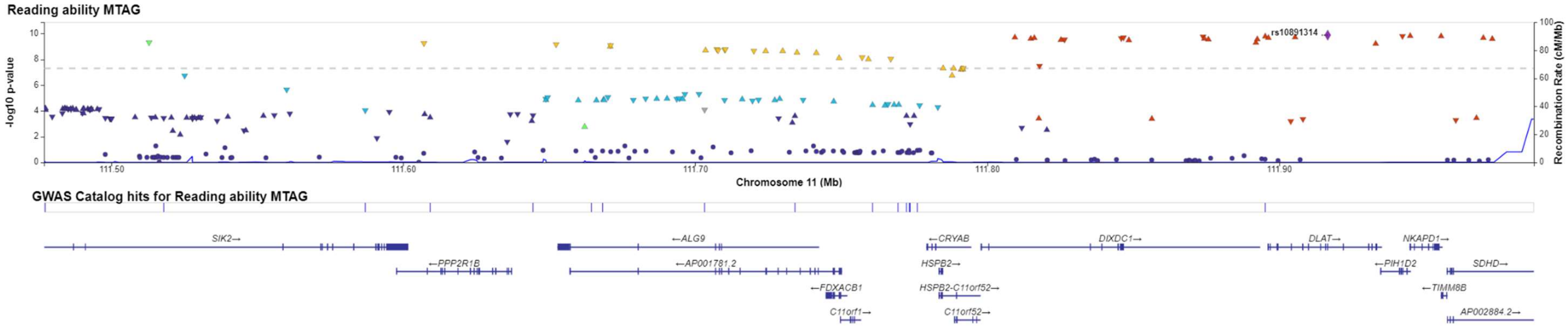


**Supplementary figure 2-24**: LocusZoom of regions associated with reading ability, region chr12q24.12 rs1265564.

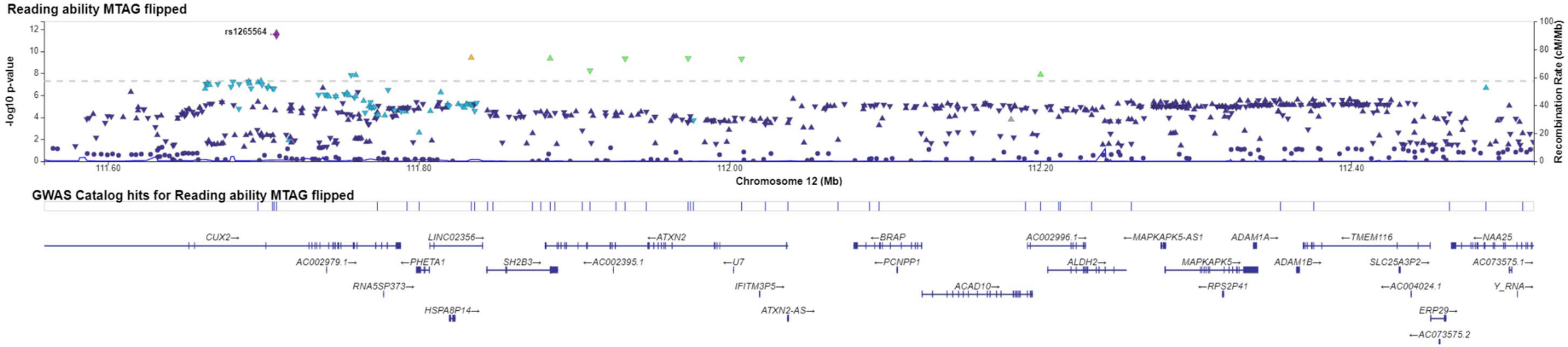

**Supplementary figure 2-25**: LocusZoom of regions associated with reading ability, region chr13q14.2 rs7996852.

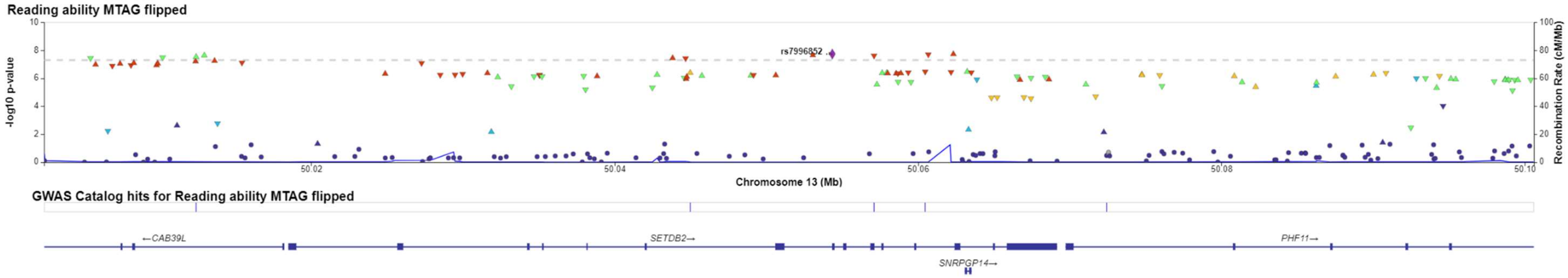


**Supplementary figure 2-26**: LocusZoom of regions associated with reading ability, region chr13q21.2 rs9538299.

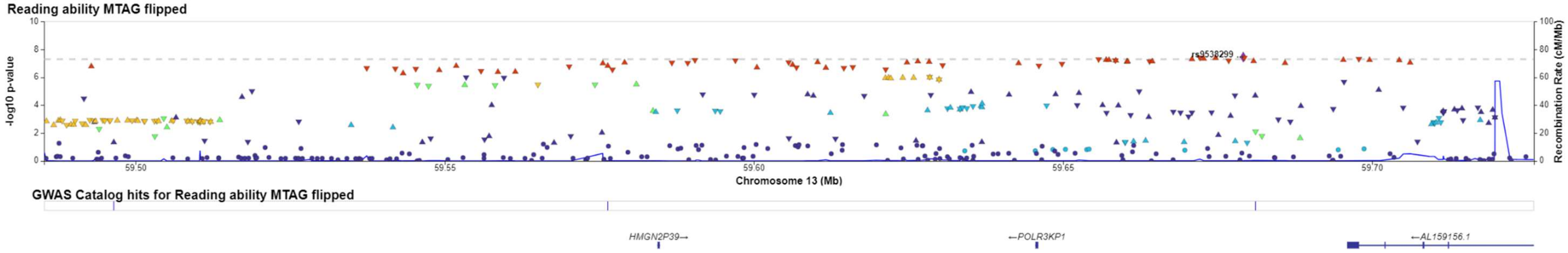

**Supplementary figure 2-27**: LocusZoom of regions associated with reading ability, region chr16p12.13 rs2015573.

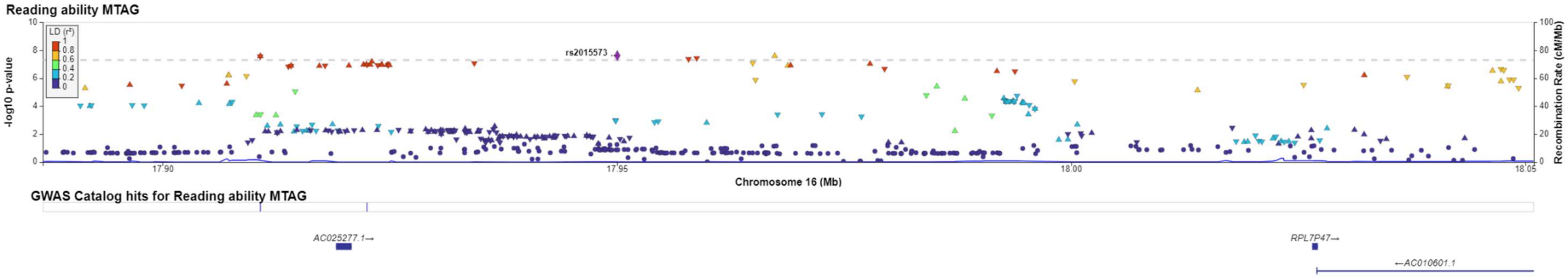


**Supplementary figure 2-28**: LocusZoom of regions associated with reading ability, region chr16q22.2 rs2158266.

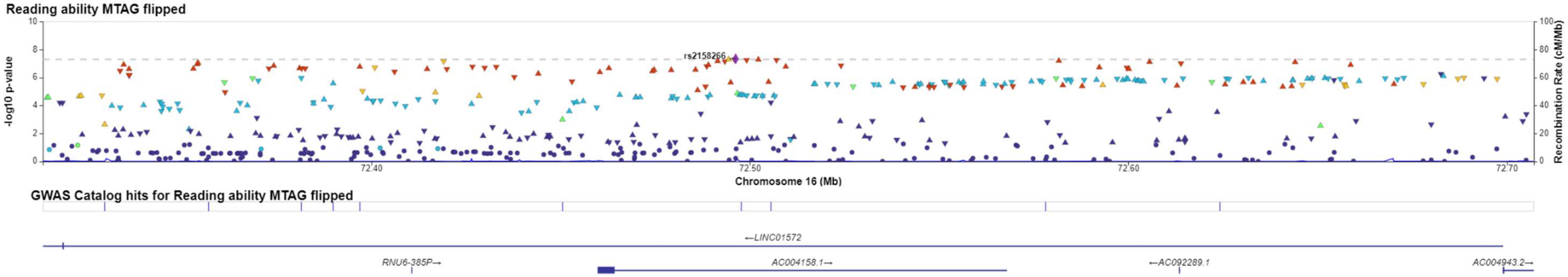

**Supplementary figure 2-29**: LocusZoom of regions associated with reading ability, region chr17q12 rs1075218.

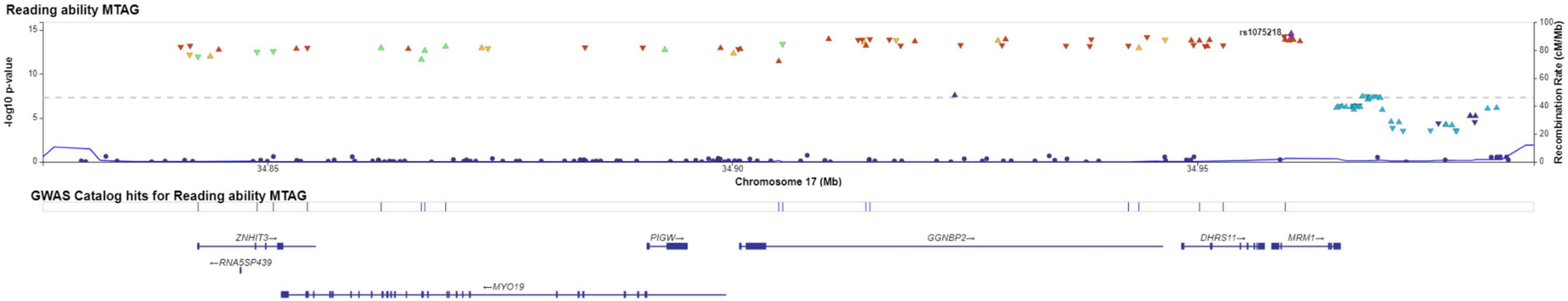


**Supplementary figure 2-30**: LocusZoom of regions associated with reading ability, region chr17q12 rs12453682.

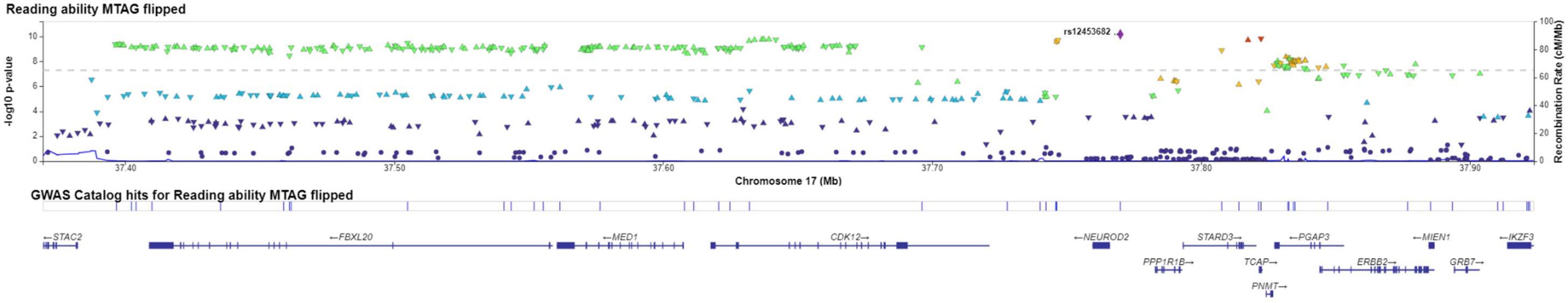

**Supplementary figure 2-31**: LocusZoom of regions associated with reading ability, region chr17q23.3 rs72841392.

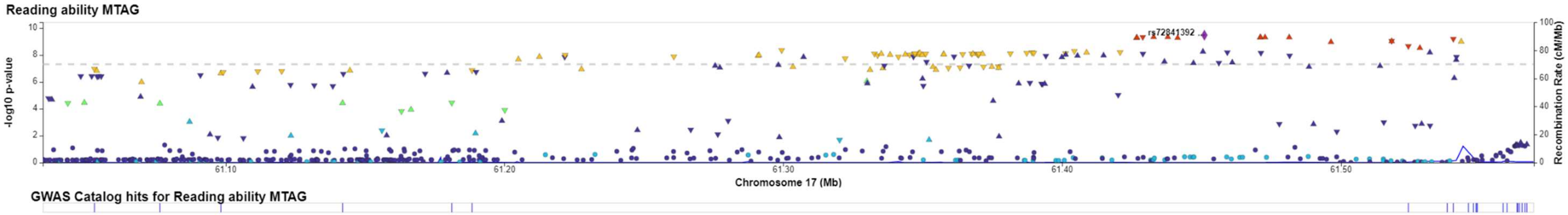


**Supplementary figure 2-32**: LocusZoom of regions associated with reading ability, region chr18q11.2 rs7505854.

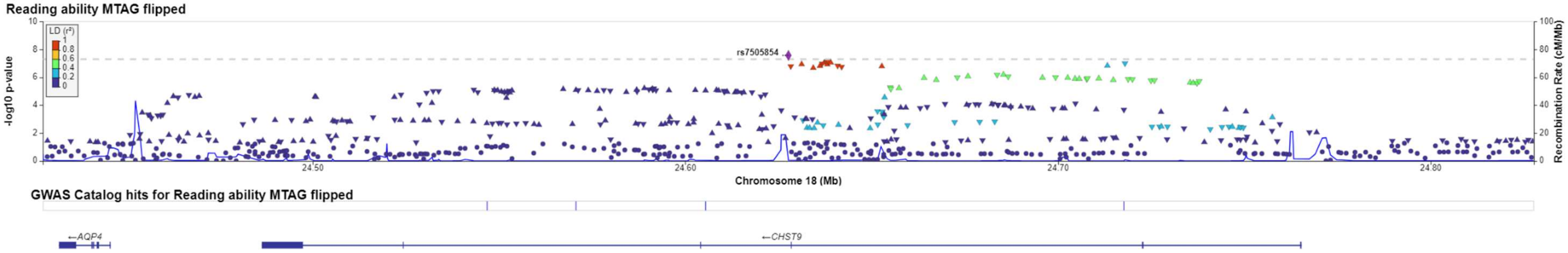

**Supplementary figure 2-33**: LocusZoom of regions associated with reading ability, region chr20q11.21 rs4911109.

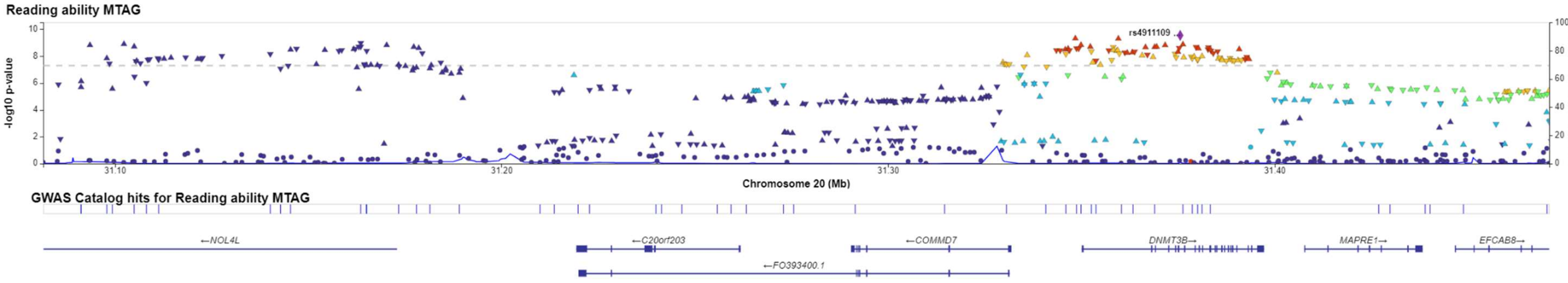


**Supplementary figure 2-34**: LocusZoom of regions associated with reading ability, region chr20q13.13 rs348258.

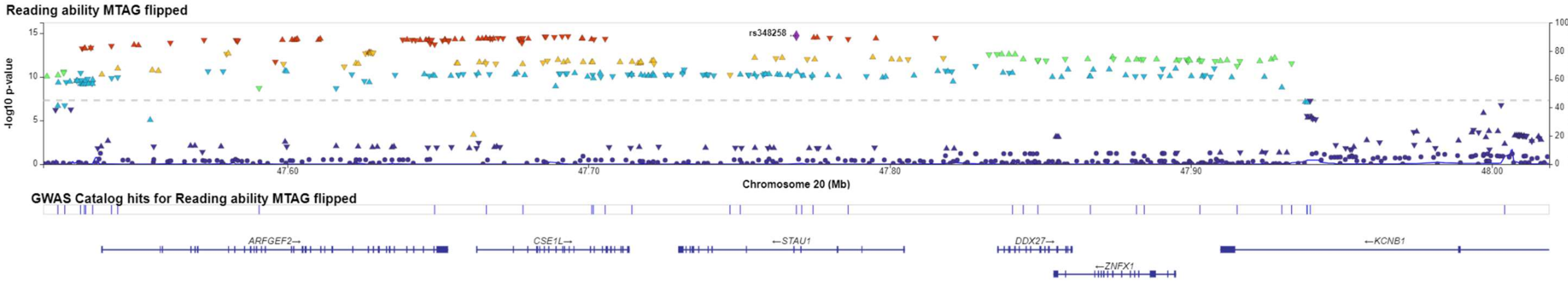

**Supplementary figure 2-35**: LocusZoom of regions associated with reading ability, region chrXq27.3 rs6626462.

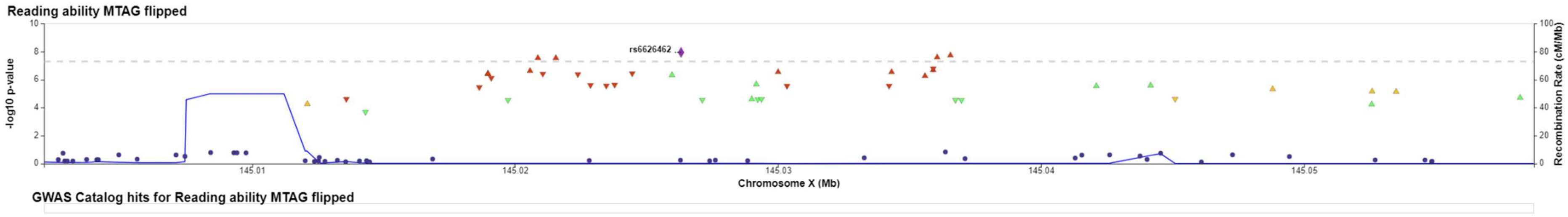

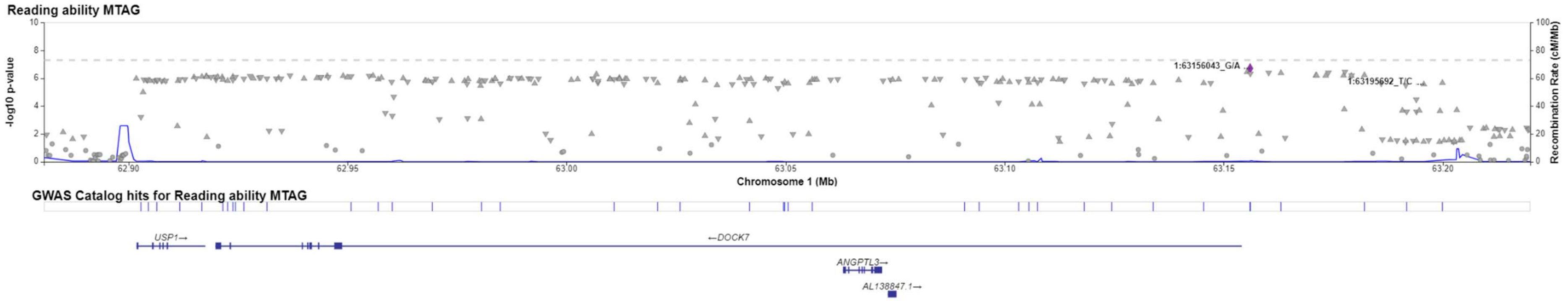


**Supplementary Figure 3:** LocusZoom of suggestive significance chr1:62900811-63199936 at P = 1.96x10$^{-7}$ with rs1168114 (1:63156043 G/A). This region overlaps completely with the locus previously reported by Eising et al 2022, however lead SNP has changed from rs11208009 to rs1168114.

A

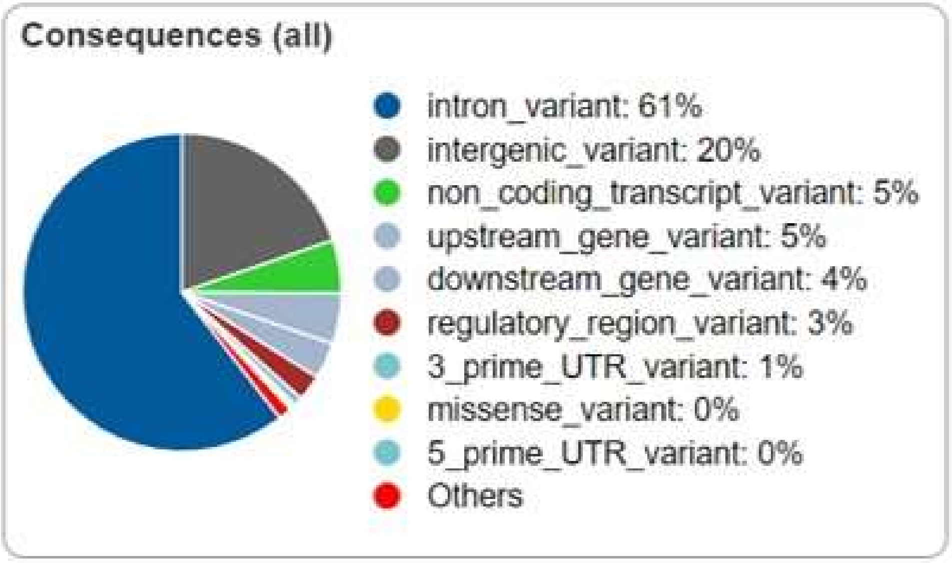


B

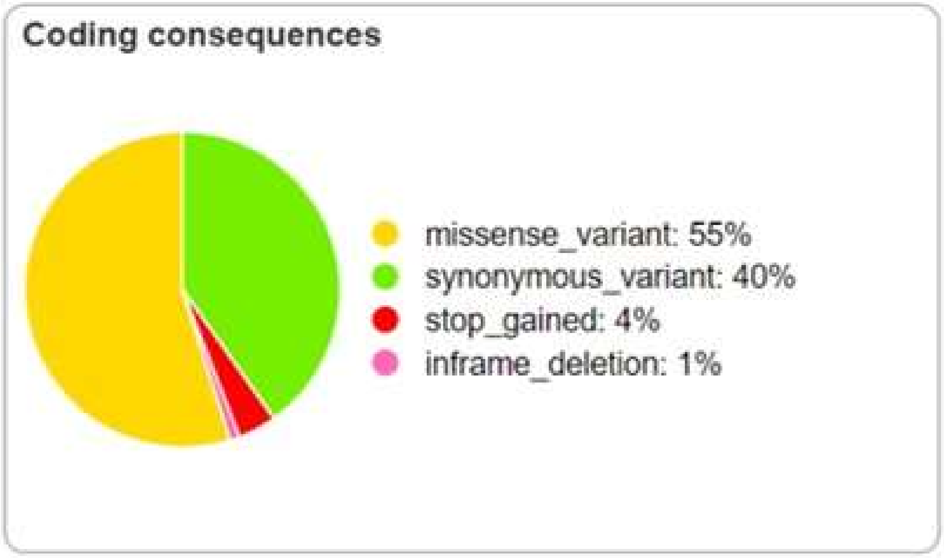


**Supplementary Figure 4:** Variant consequences annotated by Variant Effect Predictor (VEP) showing A) positional annotation of the SNP, and for the SNPs that fell within a coding region, B) the functional consequence of that variant associated with reading ability from MTAG analysis.

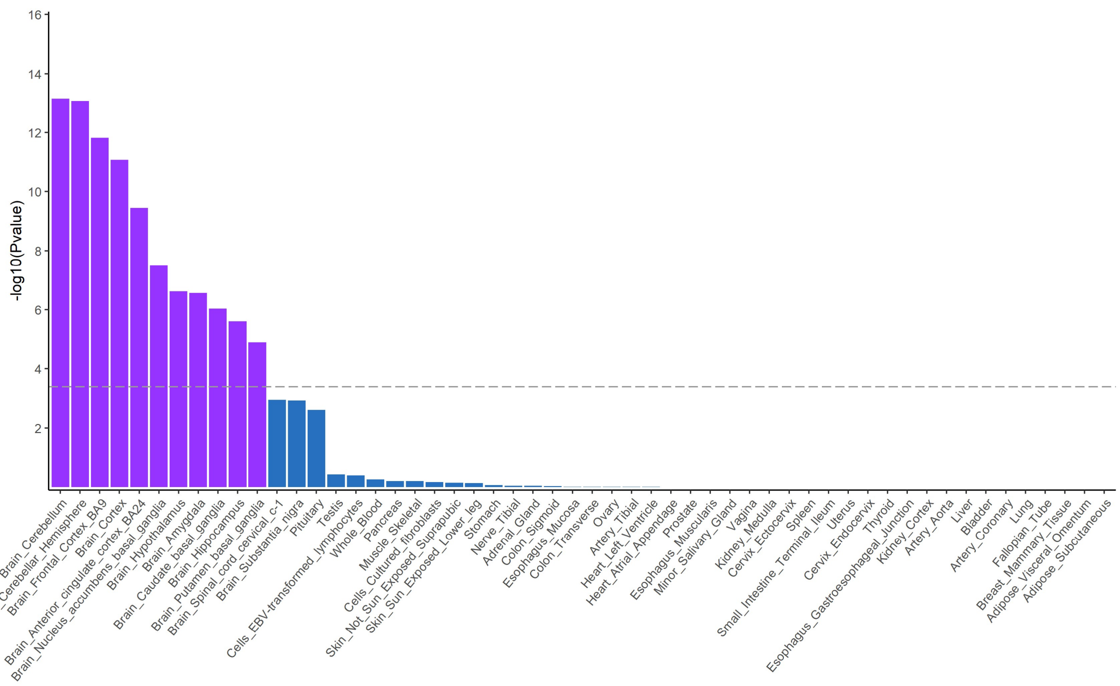


**Supplementary Figure 5:** $-\log_{10}$ P values from a MAGMA gene-property analysis of reading ability multivariate GWAS showing enrichment of expression in brain regions. The dashed line indicates the Bonferroni correction for 124 tests to $P < 4.032 \times 10^{-4}$.

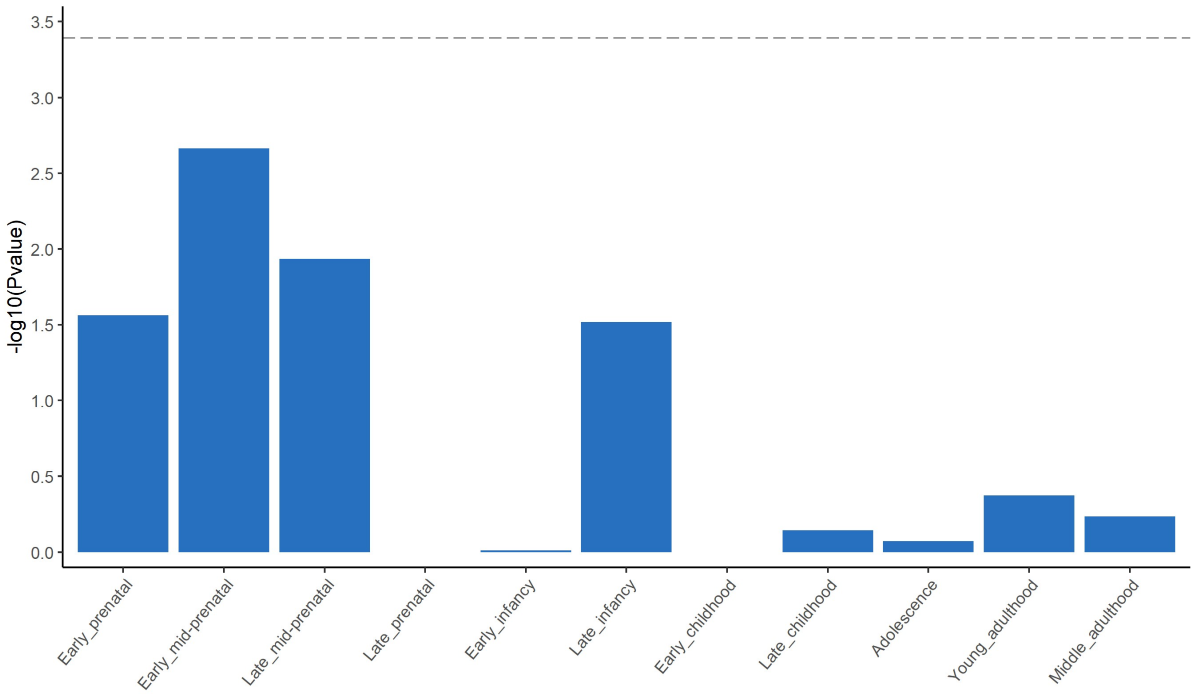


**Supplementary Figure 6:** $-\log_{10}$ P values from a MAGMA gene property analysis of reading ability multivariate GWAS showing enrichment through 11 developmental stages from BrainSpan. The dashed line indicates the Bonferroni correction for 124 tests to $P < 4.032 \times 10^{-4}$.

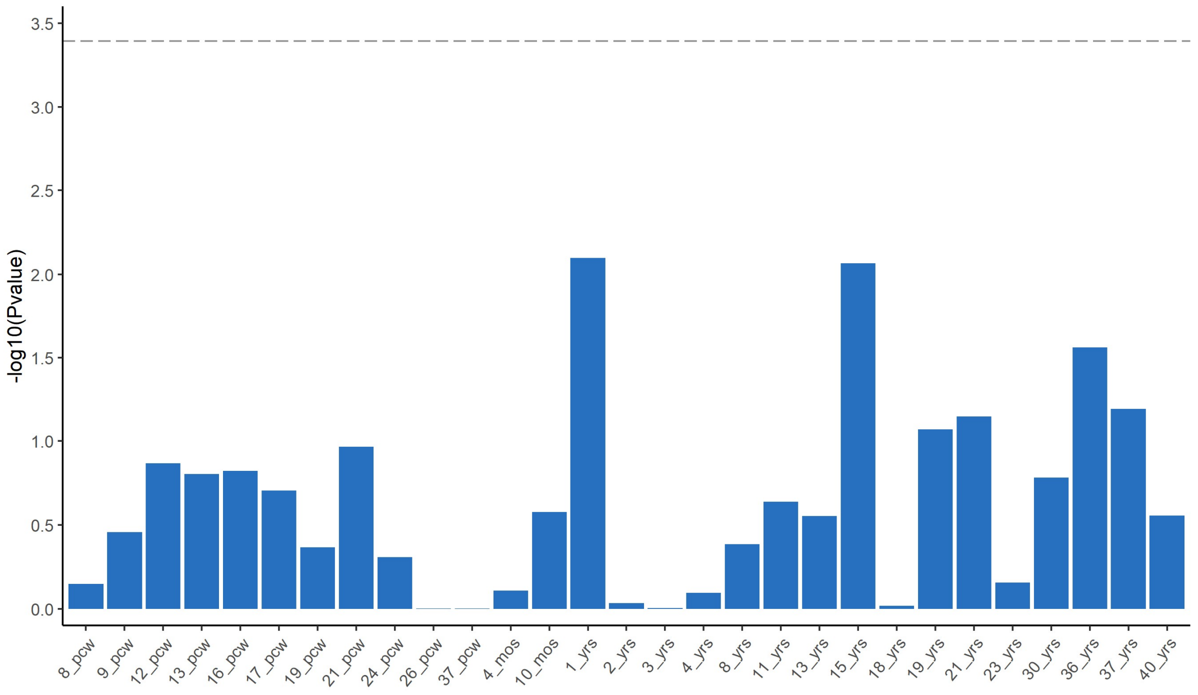


**Supplementary Figure 7:** $-\log_{10}$ P values from a MAGMA gene property analysis of reading ability multivariate GWAS showing enrichment through 29 human brain ages from BrainSpan. The dashed line indicates the Bonferroni correction for 124 tests to $P < 4.032 \times 10^{-4}$.

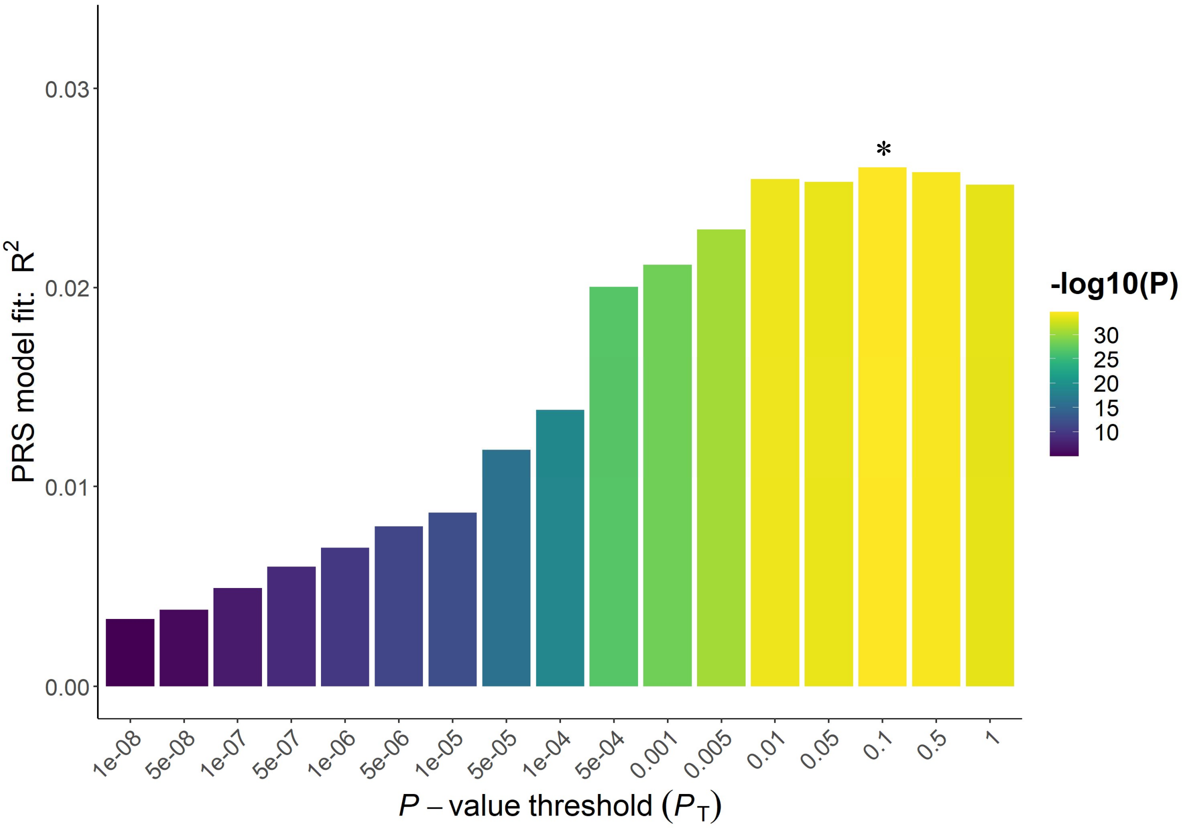


**Supplementary Figure 8:** Threshold PRS model fit for polygenic score of reading ability predicting a composite measure of reading ability at age 7 in the NCDS cohort. X axis indicated the range of P value cutoffs tested, and Y axis shows the PRS model fit ($R^2$) at each threshold. * indicates the best P value threshold for each PRS model.

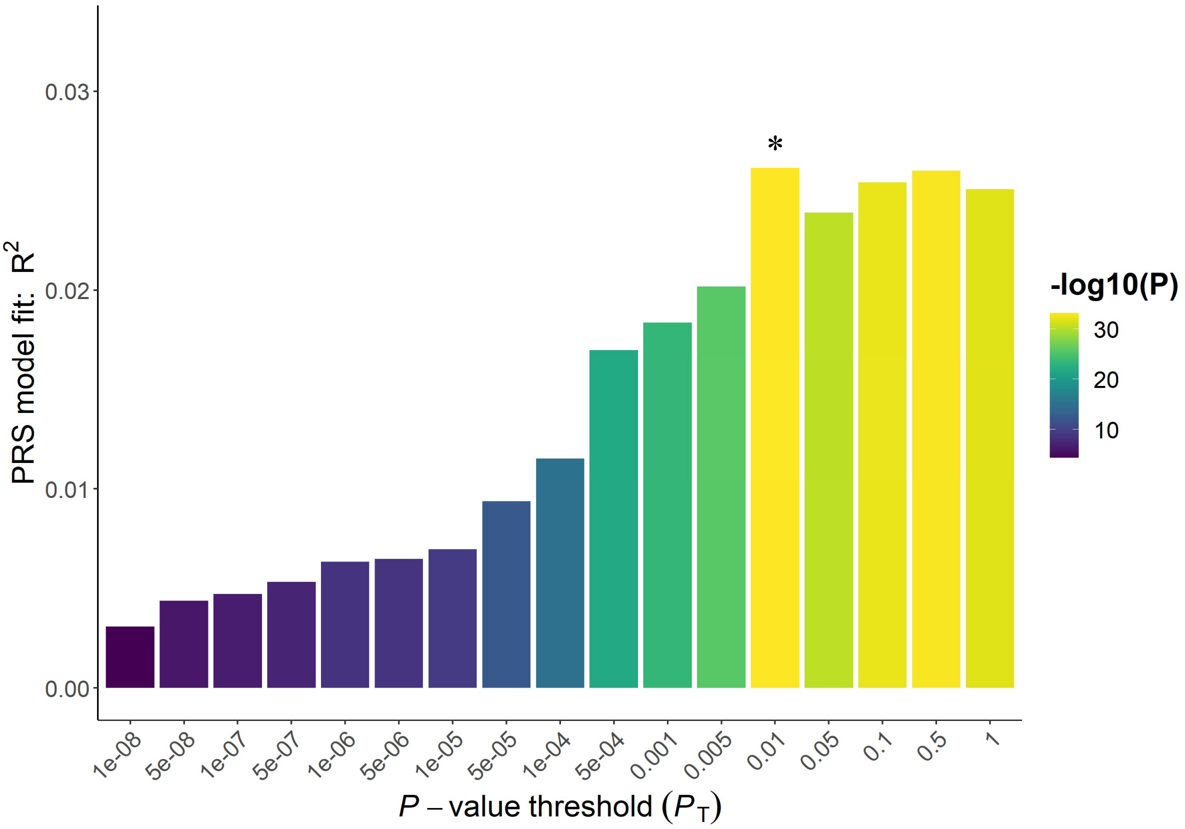


**Supplementary Figure 9:** Threshold PRS model fit for polygenic score of reading ability predicting a composite measure of reading ability at age 11 in the NCDS cohort. X axis indicated the range of P value cutoffs tested, and Y axis shows the PRS model fit ($R^2$) at each threshold.

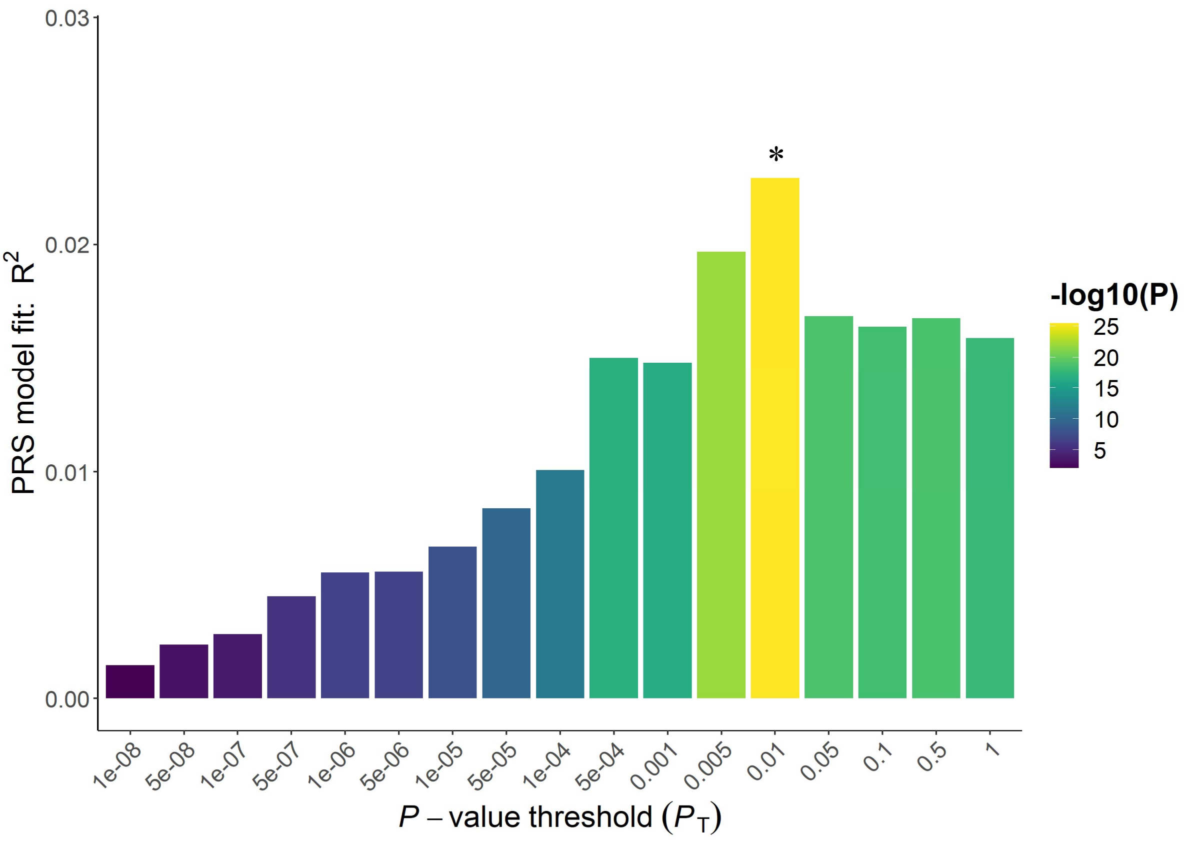


**Supplementary Figure 10:** Threshold PRS model fit for polygenic score of reading ability predicting a composite measure of reading ability at age 16 in the NCDS cohort. X axis indicated the range of P value cutoffs tested, and Y axis shows the PRS model fit ($R^2$) at each threshold.

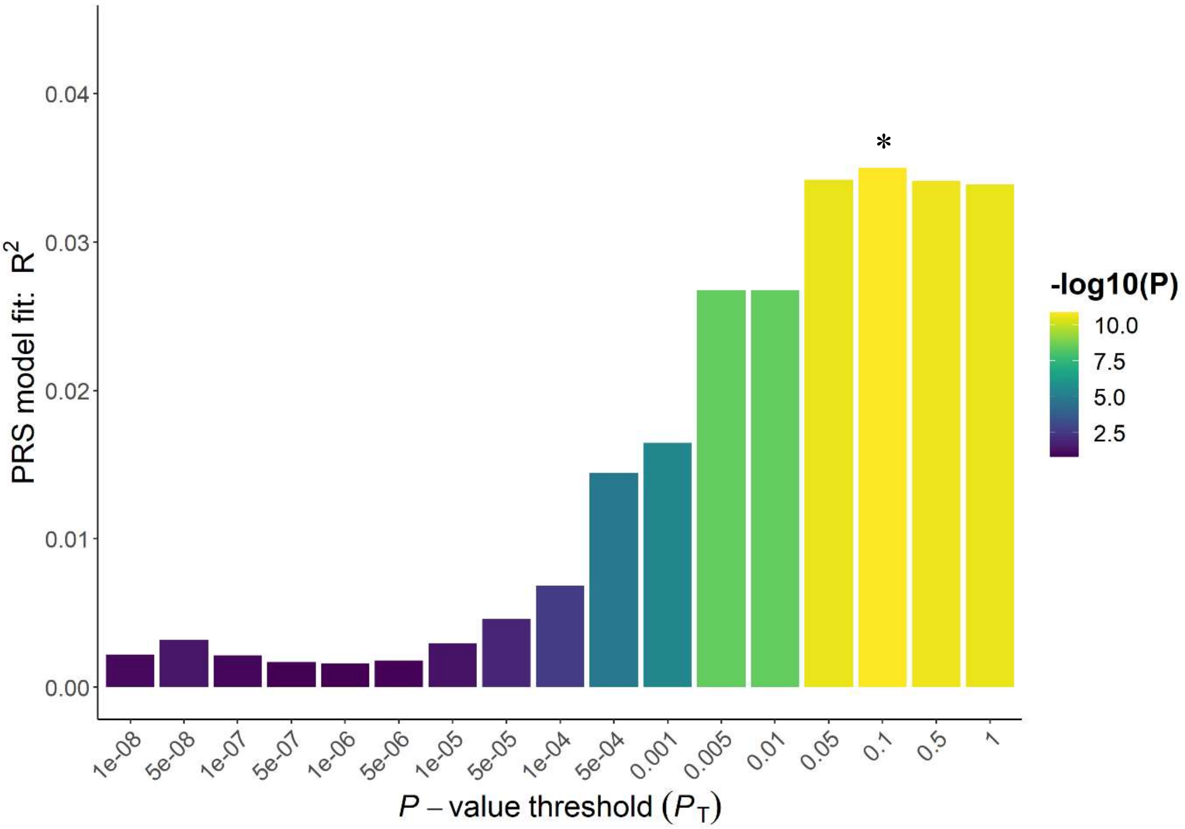


**Supplementary Figure 11:** Threshold PRS model fit for polygenic score of reading ability predicting a binary measure reading ability at age 23 in the NCDS cohort. X axis indicated the range of P value cutoffs tested, and Y axis shows the PRS model fit ($R^2$) at each threshold.

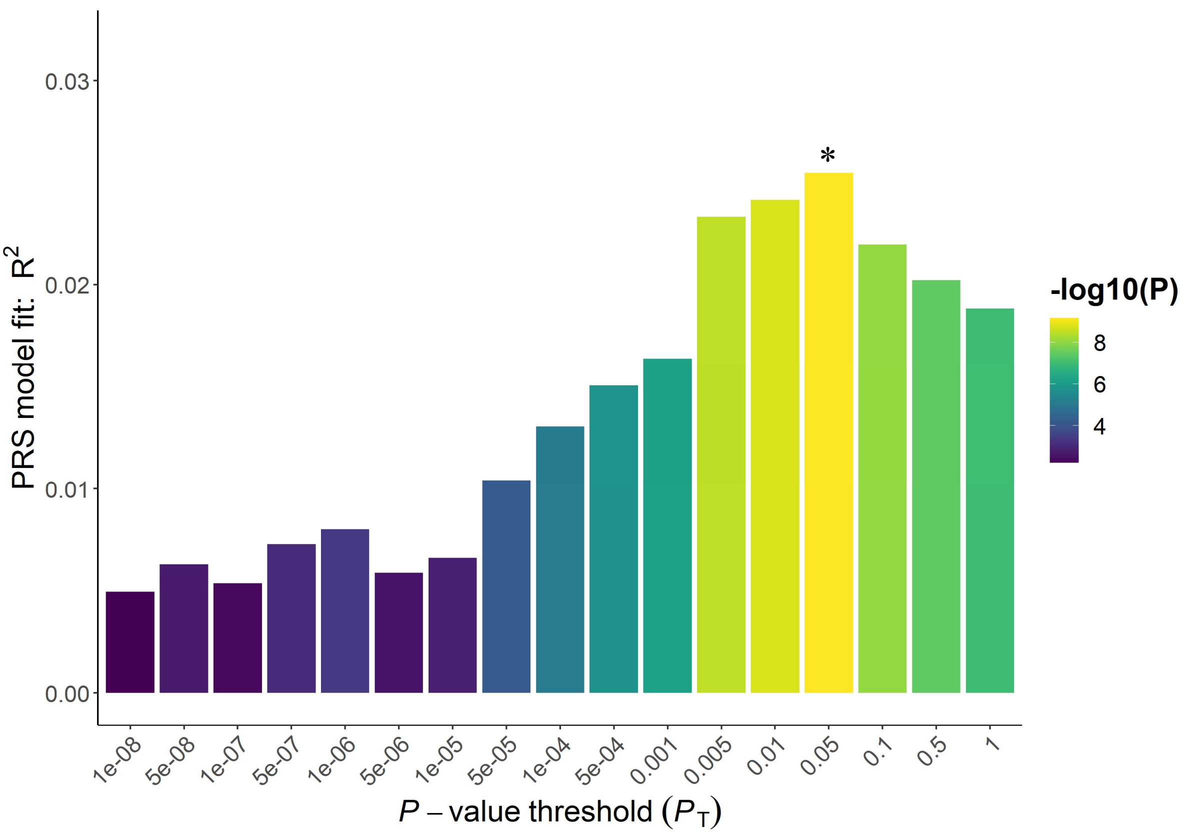


**Supplementary Figure 12:** Threshold PRS model fit for polygenic score of reading ability predicting a binary measure of reading ability at age 33 in the NCDS cohort. X axis indicated the range of P value cutoffs tested, and Y axis shows the PRS model fit ($R^2$) at each threshold.

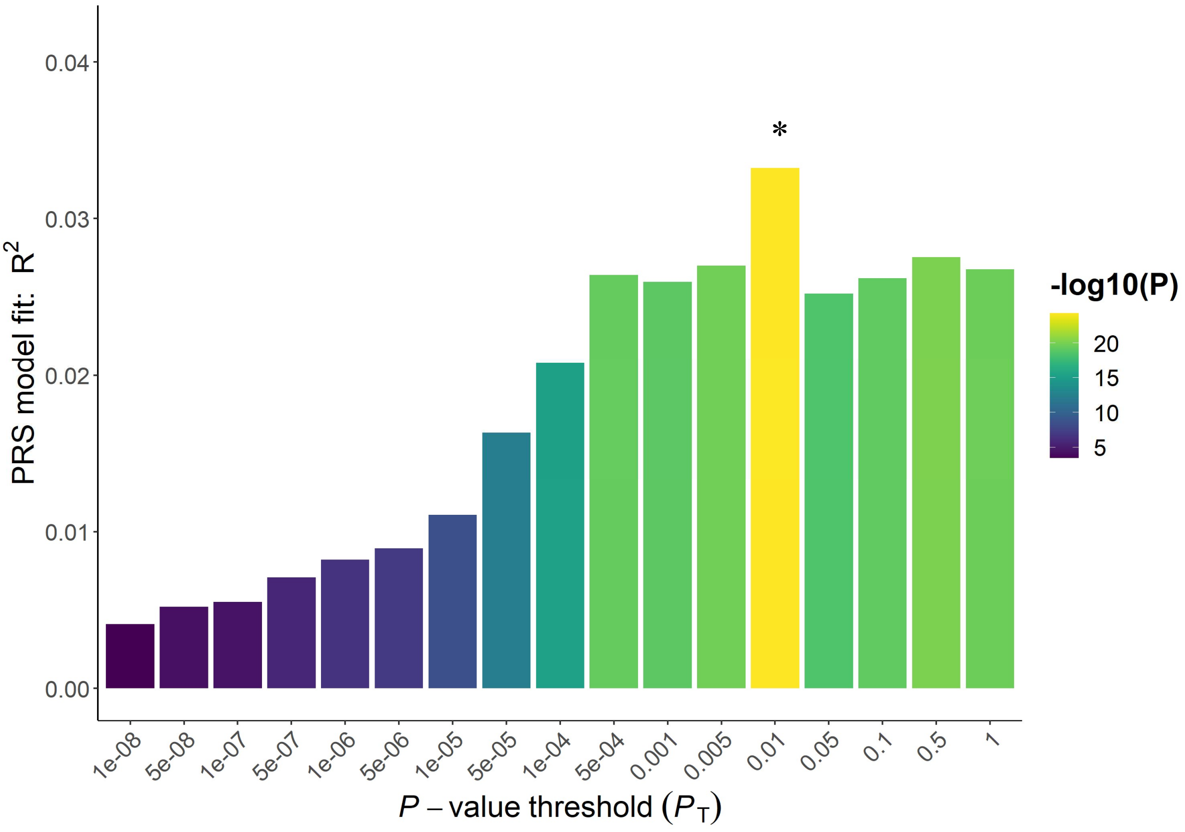


**Supplementary Figure 13:** Threshold PRS model fit for polygenic score of reading ability predicting a composite measure of reading ability across all ages in the NCDS cohort. X axis indicated the range of P value cutoffs tested, and Y axis shows the PRS model fit ($R^2$) at each threshold.
